# Mortality with concurrent treatment with gabapentin and opioids among people with spine diagnoses in the U.S. Medicare population: a propensity-matched cohort study

**DOI:** 10.1101/2024.04.26.24306460

**Authors:** Laura S. Gold, Patrick J. Heagerty, Ryan N. Hansen, Janna L. Friedly, Sandra K. Johnston, Richard A. Deyo, Michele Curatolo, Judith A. Turner, Sean D. Rundell, Katherine Wysham, Jeffrey G. Jarvik, Pradeep Suri

## Abstract

**Importance:** Given the negative impact of opioid use on population health, prescriptions for alternative pain-relieving medications, including gabapentin, have increased. Concurrent gabapentin and opioid prescriptions are commonly reported in retrospective studies of opioid-related overdose deaths.

**Objective:** To determine whether people who filled gabapentin and opioid prescriptions concurrently (“gabapentin + opioids”) had greater mortality than those who filled an active control medication (tricyclic antidepressants [TCAs] or duloxetine) and opioids concurrently (“TCAs/duloxetine + opioids”). We hypothesized that people treated with gabapentin + opioids would have higher mortality rates compared to people treated with TCAs/duloxetine + opioids.

**Design:** Propensity score-matched cohort study with an incident user, active control design. The median (maximum) follow-up was 45 (1093) days.

**Setting:** Population-based.

**Participants:** Medicare beneficiaries with spine-related diagnoses 2017-2019. The primary analysis included those who concurrently (within 30 days) filled ≥1 incident gabapentin + ≥1 opioid or ≥1 incident TCA/duloxetine + ≥1 opioid.

**Exposures:** People treated with gabapentin + opioids (n=67,133) were matched on demographic and clinical factors in a 1:1 ratio to people treated with TCAs/duloxetine + opioids (n=67,133).

**Main Outcomes and Measures:** The primary outcome was mortality at any time. A secondary outcome was occurrence of a major medical complication at any time.

**Results:** Among 134,266 participants (median age 73.4 years; 66.7% female), 2360 died before the end of follow-up. No difference in mortality was observed between groups (adjusted hazard ratio (HR) and 95% confidence interval (CI) for gabapentin + opioids was 0.98 (0.90, 1.06); p=0.63). However, people treated with gabapentin + opioids were at slightly increased risk of a major medical complication (1.02 (1.00, 1.04); p=0.03) compared to those treated with TCAs/duloxetine + opioids. Results were similar in analyses (a) restricted to ≤30-day follow-up and (b) that required ≥2 fills of each prescription.

**Conclusions and Relevance:** When treating pain in older adults taking opioids, the addition of gabapentin did not increase mortality risk relative to addition of TCAs or duloxetine. However, providers should be cognizant of a small increased risk of major medical complications among opioid users initiating gabapentin compared to those initiating TCAs or duloxetine.

## Introduction

Given increased knowledge of the negative impact of opioid use, many clinicians have turned to alternative pain management strategies. United States (US) pharmacy claims data showed decreasing numbers of opioid prescriptions from 2013 to 2018, alongside an increase in gabapentin use and concurrent opioid and gabapentin use.^1^ Although deaths due to gabapentin alone are rare, concurrent exposures to gabapentin and opioids have been noted in a high proportion of opioid-related overdose deaths.^1-3^ In 2019, the Food and Drug Administration warned^4^ that gabapentin may cause serious breathing difficulties in those with respiratory risk factors such as concurrent use of central nervous system depressants (e.g., opioids) or in older adults.^5^

Though this is concerning given continued high rates of opioid-related mortality in the US,^6^ most reports to date have not accounted for a wide range of factors that may confound the gabapentin-mortality relationship among opioid users.^1-4^ For instance, people prescribed opioids and gabapentin concurrently may have a higher prevalence of factors that predispose them to mortality compared to people using opioids alone, including the pain conditions for which they are being treated, other pain conditions, mental health conditions, and other comorbidities, compared to those taking comparator treatments. Additionally, most prior studies have not used an active control group,^7-9^ which can reduce confounding related to the need for an additional pain medication beyond opioids alone, or have not taken steps to ensure that opioids and gabapentin were used concurrently,^10^ which is relevant since the theorized mechanism of harm is the *interaction* of gabapentin with opioids.

The purpose of this research was to evaluate mortality in US Medicare beneficiaries with spine-related conditions prescribed gabapentin and opioids concurrently (“gabapentin + opioids”) and those prescribed an active control medication (tricyclic antidepressants [TCAs] or duloxetine) and opioids concurrently (“TCAs/duloxetine + opioids”). TCAs and duloxetine were chosen as active control medications because, like gabapentin, they are commonly prescribed to treat spinal and neuropathic pain.^11^ We hypothesized that people treated with gabapentin + opioids would have higher mortality rates compared to people treated with TCAs/duloxetine + opioids.

## Methods

A complete description of the study methods and analytic plan was posted on Open Science Framework (https://osf.io/9u6re/)^12^ before analyses began. The analysis plan and reporting in this manuscript adhere to RECORD-PE^13^ and START-RWE^14^ guidelines. This study was determined to be exempt from review by the University of Washington Institutional Review Board.

### Study Population

We identified enrollees from the 100% sample of Centers for Medicare and Medicaid Services (CMS) beneficiaries who had International Classification of Diseases, 10th Revision, Clinical Modification (ICD-10-CM) diagnosis codes^15^ from 2017-2019 for spine-related conditions^16-18^ (e.g., neck pain, low back pain, spinal stenosis, radiculopathy, others; eTable 1). In this article, we use the terms “prescriptions” and “fills” to refer to prescriptions that were *filled*. We excluded individuals with the following on or before an “index” fill date: age <65 years; not enrolled in CMS Part A, Part B and Part D for ≥1 year before index; enrolled in a health maintenance organization (HMO) health plan; or “red flag” spine conditions (e.g., fracture) or seizure disorders (to exclude people using gabapentin for primary seizure disorder; eTable 2).

### Medication Exposure Groups

Medication exposures were ascertained from Medicare Part D claims. People with incident gabapentin fills (no gabapentin fills during the previous year) (eTable 3) and concurrent (within 30 days) opioid fills were compared to an active control group of people who had incident TCA or duloxetine fills (no TCA/duloxetine fills in prior year) and concurrent (within 30 days) opioid fills (eFigures 1a-1b). Selective serotonin reuptake inhibitors and serotonin-noradrenaline reuptake inhibitors (other than duloxetine) were not included in the active control group because they are less effective and less commonly prescribed for pain.^19^ To ensure that opioids and the comparator treatment were likely to have been taken concurrently, all subjects had an “index fill” and a “qualifying fill.” For people in the gabapentin + opioids group, (1) the index fill could have been the calendar date of an opioid fill and the qualifying fill was the date on which they began gabapentin, or (2) the index fill could have been the calendar date on which they began gabapentin and the qualifying fill was the date of their opioid fill. Similarly, for people in the concurrent opioid-TCA/duloxetine group, (1) the index fill could have been the calendar date on which a person filled an opioid and the qualifying fill was the date on which they began TCA/duloxetine, or (2) the index fill could have been the calendar date on which they began TCA/duloxetine and the qualifying fill was the date of their opioid fill.

The primary analysis examined people who filled ≥1 gabapentin + ≥1 opioid within a 30-day period compared to those who filled ≥1 TCA/duloxetine + ≥1 opioid within 30 days. This analysis was designed to capture early mortality based on the putative mechanism of risk with concurrent treatment with opioids and gabapentin.^4^ We also conducted a secondary analysis designed to increase the likelihood that people actually consumed the medications that they filled. The typical refill period for gabapentin is 1-3 months,^20^ so this secondary analysis required ≥2 gabapentin fills + ≥2 opioid fills within a 120-day exposure period and compared these people to those with ≥2 TCA/duloxetine fills + ≥2 opioid fills in 120 days (eFigures 1c-1d).

### Censoring Variables and Outcomes

Beginning from the day of the qualifying fill, we assessed mortality as the primary outcome. The occurrence of a major medical complication (eTable4) was a secondary outcome designed to capture non-fatal sequelae related to possible side effects of gabapentin and/or TCA/duloxetine involving primarily the respiratory^21-23^ and cardiovascular^24,25^ organ systems, and other diagnoses that we had previously found to be independently predictive of mortality in our prior work in the Medicare population (eTable 4).^26^ We also examined early mortality and complications at 30 days. Censoring events were fills of benzodiazepine or pregabalin after the qualifying fill (because concurrent use of benzodiazepines independently increases mortality among opioid users^27^ and pregabalin is pharmacologically similar to gabapentin^28^); no opioid fill for >45 days; TCA/duloxetine fill in comparison groups treated with gabapentin; no gabapentin fill for >180 days in comparison groups treated with gabapentin; gabapentin fill in comparison groups treated with TCA/duloxetine; no TCA/duloxetine fill for >180 days in comparison groups treated with TCA/duloxetine; or the end of the study (December 31, 2019), whichever occurred first.

### Statistical Analysis

To ensure internal validity and limit confounding, we used two approaches to ensure that the TCA/duloxetine group was as comparable as possible to the gabapentin group in terms of covariates that were potentially predictive of mortality. First, we included a wide range of variables in the logistic regression used for propensity score matching (PSM) based on clinical knowledge and prior literature: age, sex, race, region, indicator of state buy-in (a surrogate for socioeconomic status), time from first spine-related diagnosis code to index, index year, Charlson co-morbidity score,^29^ diagnoses in the year prior to index (non-skin cancer, metastatic cancer, congestive heart failure, dementia, weight loss, hemiplegia, alcohol abuse, cardiac arrhythmias, chronic pulmonary disease, coagulopathy, complicated diabetes, deficiency anemia, fluid/electrolyte disorders, liver disease, peripheral vascular disease, mood disorders, pulmonary circulation disorders, HIV/AIDS, hypertension, myocardial infarction, rheumatic disease, peptic ulcer disease, diabetes without complications, opioid use disorder, Parkinson disease, obesity, insomnia, sleep apnea, renal failure, spine-related neuropathic pain), healthcare utilization in the year prior to index (number of inpatient admissions, outpatient encounters, number of dates with generalized anxiety disorder (GAD) diagnoses, number of dates with post-traumatic stress disorder (PTSD) diagnoses, number of dates with back pain diagnoses, number of dates with depression diagnoses, number of dates with non-spine neuropathy diagnoses,^30^ medication fills in the year prior to index (non-steroidal anti-inflammatory drugs, acetaminophen, muscle relaxants, benzodiazepines, pregabalin, ketamine, esketamine, anti-psychotics, and anti-depressants; eTable3), receipt of therapeutic repetitive transcranial magnetic stimulation, receipt of electroconvulsive therapy, and sum of the days’ supplies of all opioid fills in the year prior to index.

Second, as we expected certain variables that occurred between the index and qualifying fills to be even more strongly related to whether people received the prescriptions of interest *and* to mortality, we matched on them “exactly” in addition to matching on the propensity score. These included (1) average daily morphine milligram equivalents (MME)^31^ of opioid prescriptions filled on/between the index and qualifying fill dates, matched within 9 categories (>0 to 2.1; >2.1 to 5.3; >5.3 to 10.0; >10.0 to 15; >15 to 20; >20 to 30; >30 to 40; >40 to 64, and >64) based on MME distribution; (2) the length of time between the index and qualifying fills (to make the timing of concurrent opioid treatment comparable between groups); and (3) major medical complications that occurred between the index and qualifying fills (eTable 4).

We calculated descriptive statistics (numbers, proportions, medians, interquartile ranges (IQRs), and standardized mean differences) to ensure the matched groups were similar. Kaplan-Meier curves were created to graphically assess the proportional hazards assumption, which was met for all analyses. Cox proportional hazard regression was used to estimate adjusted hazard ratios (aHRs) and 95% confidence intervals (95% CIs). All regression models were adjusted for age, race, sex, geographical location, year of index fill, Charlson comorbidity score, filling prescription pain medications prior to index, and the number of inpatient and outpatient encounters in the year prior to index. Two-sided p-values of <0.05 were considered statistically significant and analyses were performed using SAS Enterprise Guide version 7.15 and SAS for Windows version 9.4, SAS Institute Inc., Cary, NC.

After we had posted our analysis plan (https://osf.io/9u6re/), Corriere et al.^10^ published a study that investigated differences in mortality between Medicare beneficiaries who filled gabapentin vs. those who filled duloxetine and reported that people in a high MME category (≥50 MME) who filled gabapentin were at twice the risk of mortality compared to those who filled duloxetine. Thus, we conducted *post hoc* analyses in which we emulated aspects of their methods, including (a) using an active control group of people who filled duloxetine only; (b) applying (a) plus *not* matching on the length of time from index to qualifying fill dates or complications from index to qualifying fill dates; and (c) applying (a) and (b) plus applying exclusion criteria used by Corriere et al.^10^ (e.g., excluding people in long-term care, with serious illnesses, age >90 years; see eFigure 2).

## Results

The flow of Medicare beneficiaries in the primary analysis is shown in Figure 1. An abbreviated list of cohort characteristics is shown in Table 1 and a complete list is shown in eTable 5. The unmatched population included 426,216 people treated with gabapentin + opioids who were eligible to be matched to 99,668 people treated with TCA/duloxetine + opioids. Of these, 67,133 people were included in each matched treatment group, which exceed the minimum sample size (n=1275 per group) estimated in power calculations we conducted *a priori*.^12^ The median (range) follow-up time was 45 (1-1093) days. Although the unmatched treatment groups differed with respect to various characteristics (eTable 5), the matched groups were closely comparable (Table 1 and eTable 5); all subsequent results pertain to the matched groups. Subjects were predominantly female and White, and about half had spine-related neuropathic pain diagnoses in the year prior to index. In both treatment groups, fills of other medications for pain and mental health diagnoses were common and the median (IQR) of the average daily MME was 18.8 (6.3, 40) between their index and qualifying dates.

**Table 1.**
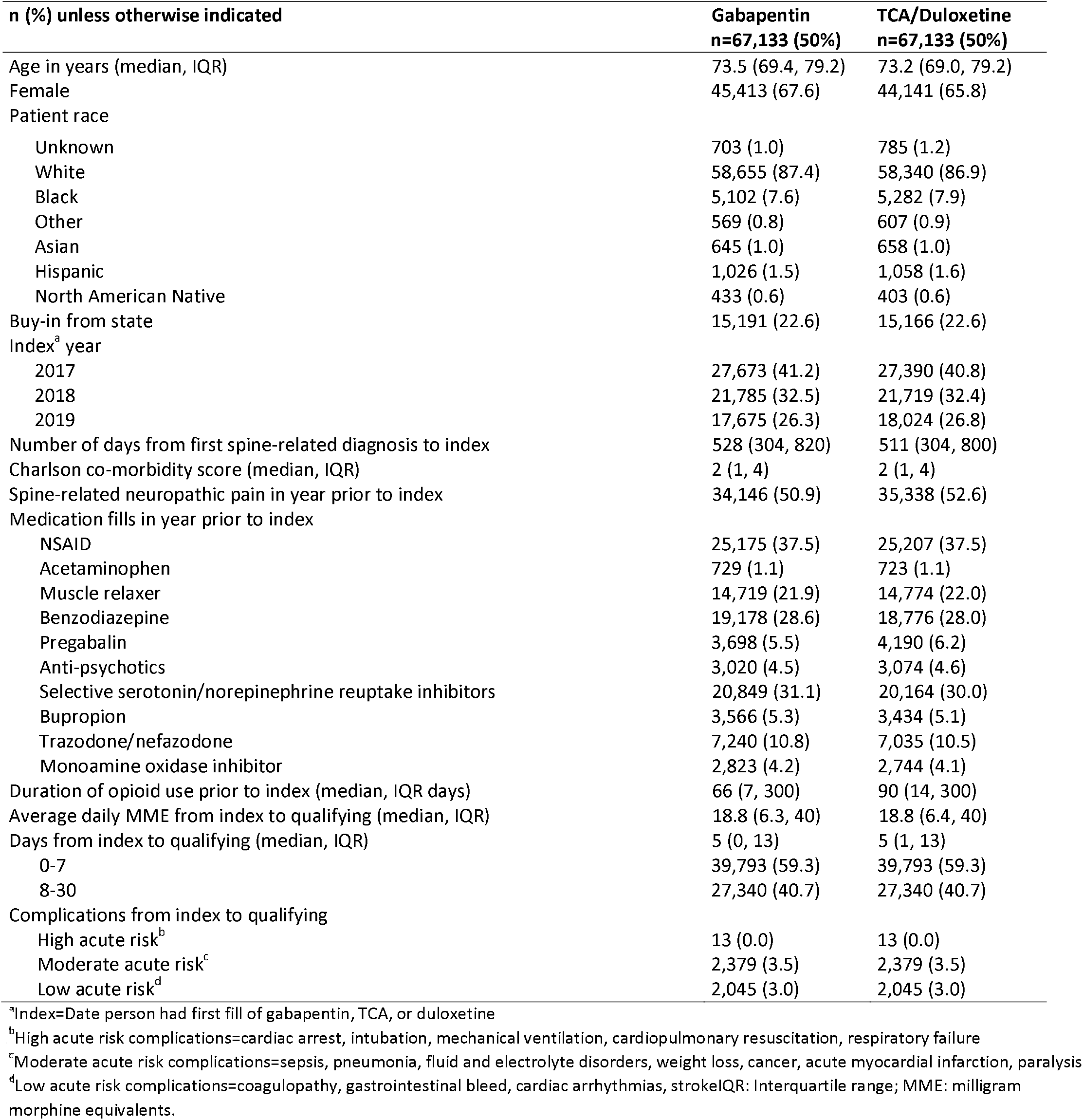
Comparison of 1:1 matched patients who received gabapentin to those who received TCAs/duloxetine in the primary analysis (selected variables).

| **n (%) unless otherwise indicated** | **Gabapentin**<br>**n=67,133 (50%)** | **TCA/Duloxetine**<br>**n=67,133 (50%)** |
| --- | --- | --- |
| Age in years (median, IQR) | 73.5 (69.4, 79.2) | 73.2 (69.0, 79.2) |
| Female | 45,413 (67.6) | 44,141 (65.8) |
| Patient race |  |  |
| Unknown | 703 (1.0) | 785 (1.2) |
| White | 58,655 (87.4) | 58,340 (86.9) |
| Black | 5,102 (7.6) | 5,282 (7.9) |
| Other | 569 (0.8) | 607 (0.9) |
| Asian | 645 (1.0) | 658 (1.0) |
| Hispanic | 1,026 (1.5) | 1,058 (1.6) |
| North American Native | 433 (0.6) | 403 (0.6) |
| Buy-in from state | 15,191 (22.6) | 15,166 (22.6) |
| Index<sup>a</sup> year |  |  |
| 2017 | 27,673 (41.2) | 27,390 (40.8) |
| 2018 | 21,785 (32.5) | 21,719 (32.4) |
| 2019 | 17,675 (26.3) | 18,024 (26.8) |
| Number of days from first spine-related diagnosis to index | 528 (304, 820) | 511 (304, 800) |
| Charlson co-morbidity score (median, IQR) | 2 (1, 4) | 2 (1, 4) |
| Spine-related neuropathic pain in year prior to index | 34,146 (50.9) | 35,338 (52.6) |
| Medication fills in year prior to index |  |  |
| NSAID | 25,175 (37.5) | 25,207 (37.5) |
| Acetaminophen | 729 (1.1) | 723 (1.1) |
| Muscle relaxer | 14,719 (21.9) | 14,774 (22.0) |
| Benzodiazepine | 19,178 (28.6) | 18,776 (28.0) |
| Pregabalin | 3,698 (5.5) | 4,190 (6.2) |
| Anti-psychotics | 3,020 (4.5) | 3,074 (4.6) |
| Selective serotonin/norepinephrine reuptake inhibitors | 20,849 (31.1) | 20,164 (30.0) |
| Bupropion | 3,566 (5.3) | 3,434 (5.1) |
| Trazodone/nefazodone | 7,240 (10.8) | 7,035 (10.5) |
| Monoamine oxidase inhibitor | 2,823 (4.2) | 2,744 (4.1) |
| Duration of opioid use prior to index (median, IQR days) | 66 (7, 300) | 90 (14, 300) |
| Average daily MME from index to qualifying (median, IQR) | 18.8 (6.3, 40) | 18.8 (6.4, 40) |
| Days from index to qualifying (median, IQR) | 5 (0, 13) | 5 (1, 13) |
| 0-7 | 39,793 (59.3) | 39,793 (59.3) |
| 8-30 | 27,340 (40.7) | 27,340 (40.7) |
| Complications from index to qualifying |  |  |
| High acute risk<sup>b</sup> | 13 (0.0) | 13 (0.0) |
| Moderate acute risk<sup>c</sup> | 2,379 (3.5) | 2,379 (3.5) |
| Low acute risk<sup>d</sup> | 2,045 (3.0) | 2,045 (3.0) |

<sup>a</sup>Index=Date person had first fill of gabapentin, TCA, or duloxetine
<sup>b</sup>High acute risk complications=cardiac arrest, intubation, mechanical ventilation, cardiopulmonary resuscitation, respiratory failure
<sup>c</sup>Moderate acute risk complications=sepsis, pneumonia, fluid and electrolyte disorders, weight loss, cancer, acute myocardial infarction, paralysis
<sup>d</sup>Low acute risk complications=coagulopathy, gastrointestinal bleed, cardiac arrhythmias, stroke IQR: Interquartile range; MME: milligram
morphine equivalents.

**Figure 1.**
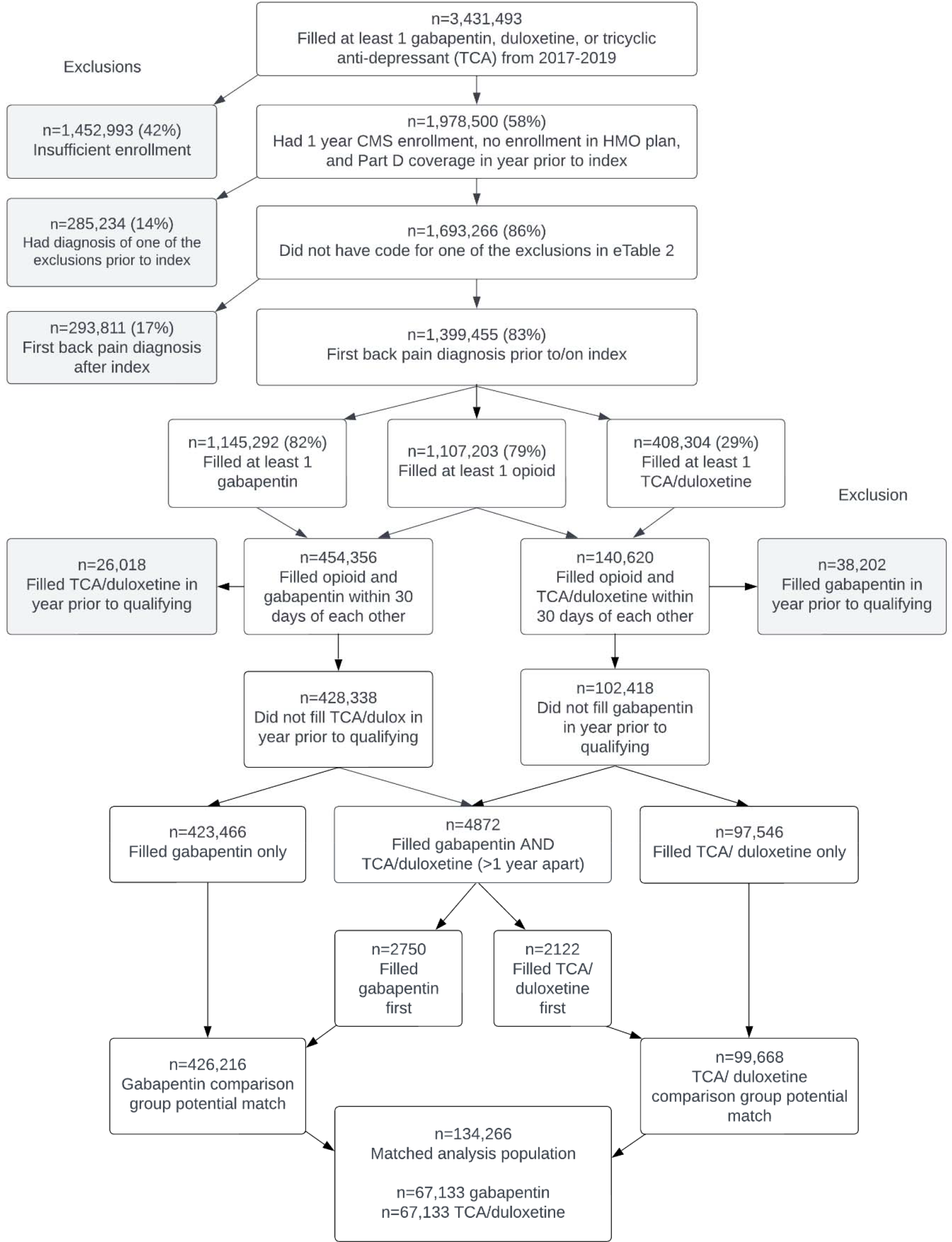
Flow of patients for the primary analysis.

The numbers and percentages of people who experienced the primary and secondary outcomes are shown in Table 2. The proportions who died were similar between the treatment groups (1.8% in the gabapentin group and 1.7% in the TCA/duloxetine group). A slightly greater proportion of people in the gabapentin group experienced a major medical complication (30.3% versus 29.1%). People in the gabapentin treatment group were somewhat more likely to have been censored because they did not refill an opioid for >45 days (54.6% versus 48.4%).

**Table 2.**
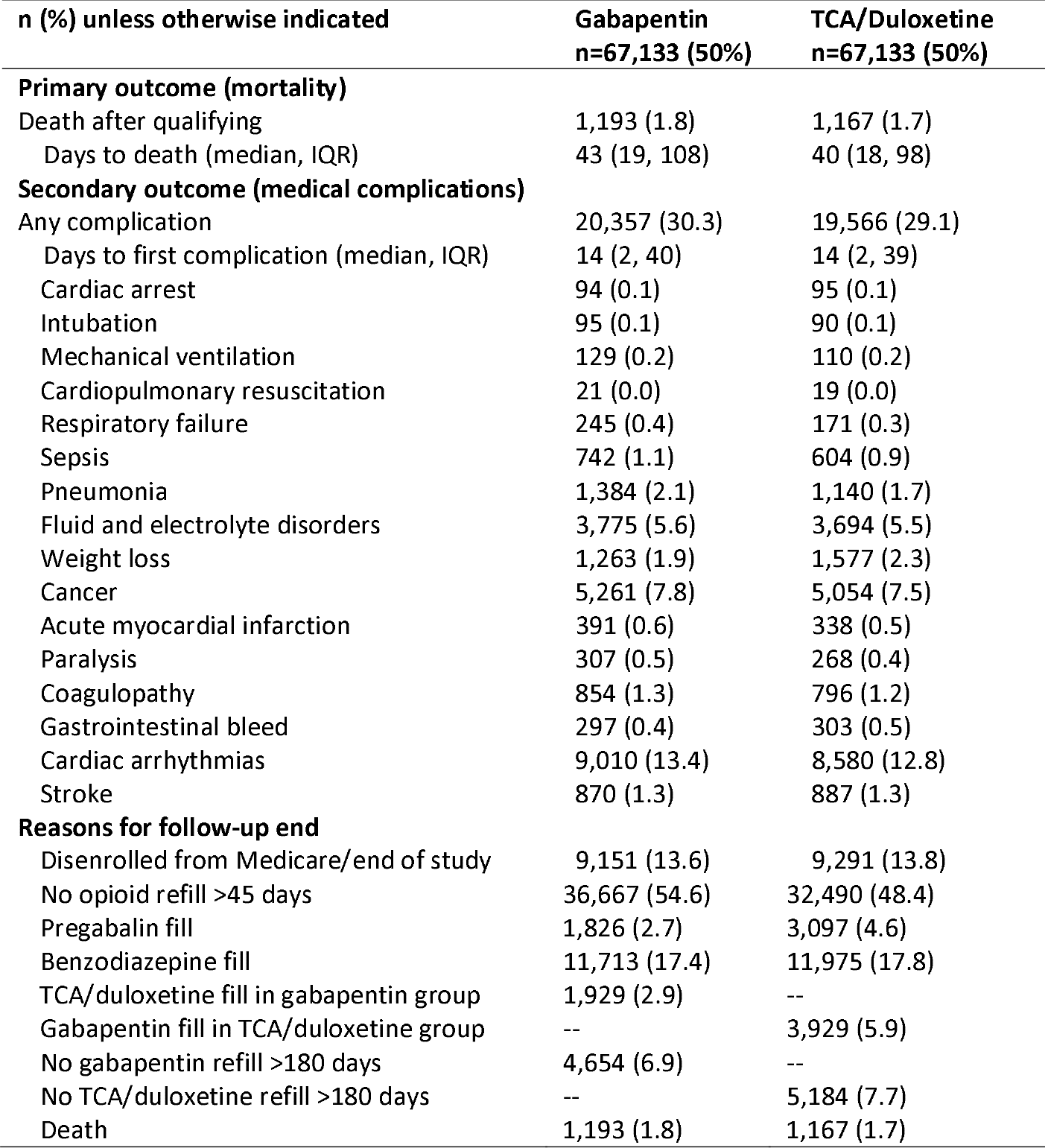
Primary analysis outcomes at any time and reasons for end of follow-up.

| n (%) unless otherwise indicated | Gabapentin<br>n=67,133 (50%) | TCA/Duloxetine<br>n=67,133 (50%) |
| --- | --- | --- |
| **Primary outcome (mortality)** |  |  |
| Death after qualifying | 1,193 (1.8) | 1,167 (1.7) |
| Days to death (median, IQR) | 43 (19, 108) | 40 (18, 98) |
| **Secondary outcome (medical complications)** |  |  |
| Any complication | 20,357 (30.3) | 19,566 (29.1) |
| Days to first complication (median, IQR) | 14 (2, 40) | 14 (2, 39) |
| Cardiac arrest | 94 (0.1) | 95 (0.1) |
| Intubation | 95 (0.1) | 90 (0.1) |
| Mechanical ventilation | 129 (0.2) | 110 (0.2) |
| Cardiopulmonary resuscitation | 21 (0.0) | 19 (0.0) |
| Respiratory failure | 245 (0.4) | 171 (0.3) |
| Sepsis | 742 (1.1) | 604 (0.9) |
| Pneumonia | 1,384 (2.1) | 1,140 (1.7) |
| Fluid and electrolyte disorders | 3,775 (5.6) | 3,694 (5.5) |
| Weight loss | 1,263 (1.9) | 1,577 (2.3) |
| Cancer | 5,261 (7.8) | 5,054 (7.5) |
| Acute myocardial infarction | 391 (0.6) | 338 (0.5) |
| Paralysis | 307 (0.5) | 268 (0.4) |
| Coagulopathy | 854 (1.3) | 796 (1.2) |
| Gastrointestinal bleed | 297 (0.4) | 303 (0.5) |
| Cardiac arrhythmias | 9,010 (13.4) | 8,580 (12.8) |
| Stroke | 870 (1.3) | 887 (1.3) |
| **Reasons for follow-up end** |  |  |
| Disenrolled from Medicare/end of study | 9,151 (13.6) | 9,291 (13.8) |
| No opioid refill >45 days | 36,667 (54.6) | 32,490 (48.4) |
| Pregabalin fill | 1,826 (2.7) | 3,097 (4.6) |
| Benzodiazepine fill | 11,713 (17.4) | 11,975 (17.8) |
| TCA/duloxetine fill in gabapentin group | 1,929 (2.9) | -- |
| Gabapentin fill in TCA/duloxetine group | -- | 3,929 (5.9) |
| No gabapentin refill >180 days | 4,654 (6.9) | -- |
| No TCA/duloxetine refill >180 days | -- | 5,184 (7.7) |
| Death | 1,193 (1.8) | 1,167 (1.7) |

We did not observe any difference in the risk of death between drug treatment groups over the entire follow-up time (eFigure 3, Figure 2) (aHR, 0.98; 95% CI, 0.90-1.06). However, people treated with gabapentin + opioids were at slight but statistically significantly greater risk of medical complications at any time compared to those treated with TCA/duloxetine + opioids (eFigures 4-5; aHR 1.02; 95% CI, 1.00-1.04). Results were similar when we examined 30-day outcomes (eTable 6; eFigures 6-9). In *post hoc* analyses to compare with work by Corriere et al.,^10^ we found no significant differences in mortality in any MME stratum (Table 3, eTable 7).

**Table 3.**
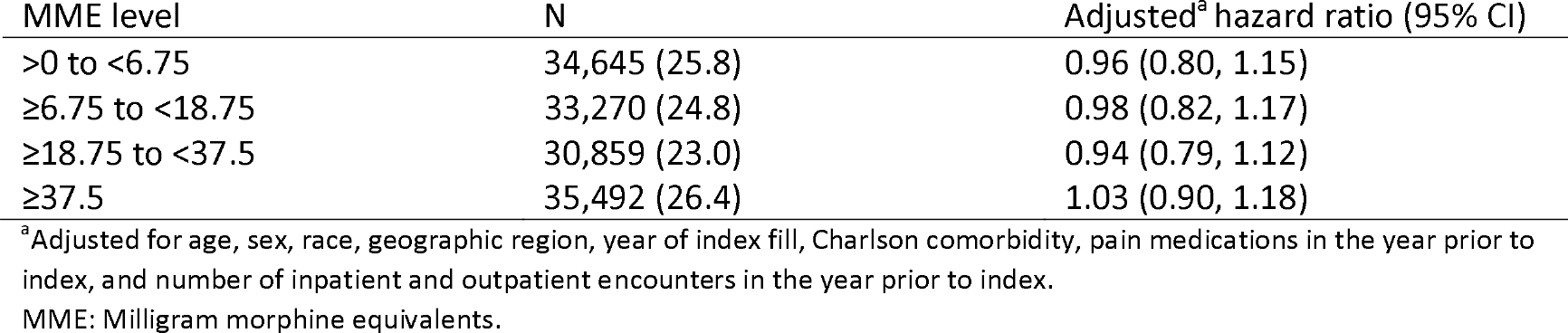
Primary analysis: *post hoc* analysis comparing hazard ratios for any-time mortality among quartiles of average daily MME from fills from index to qualifying dates.

**Figure 2.**
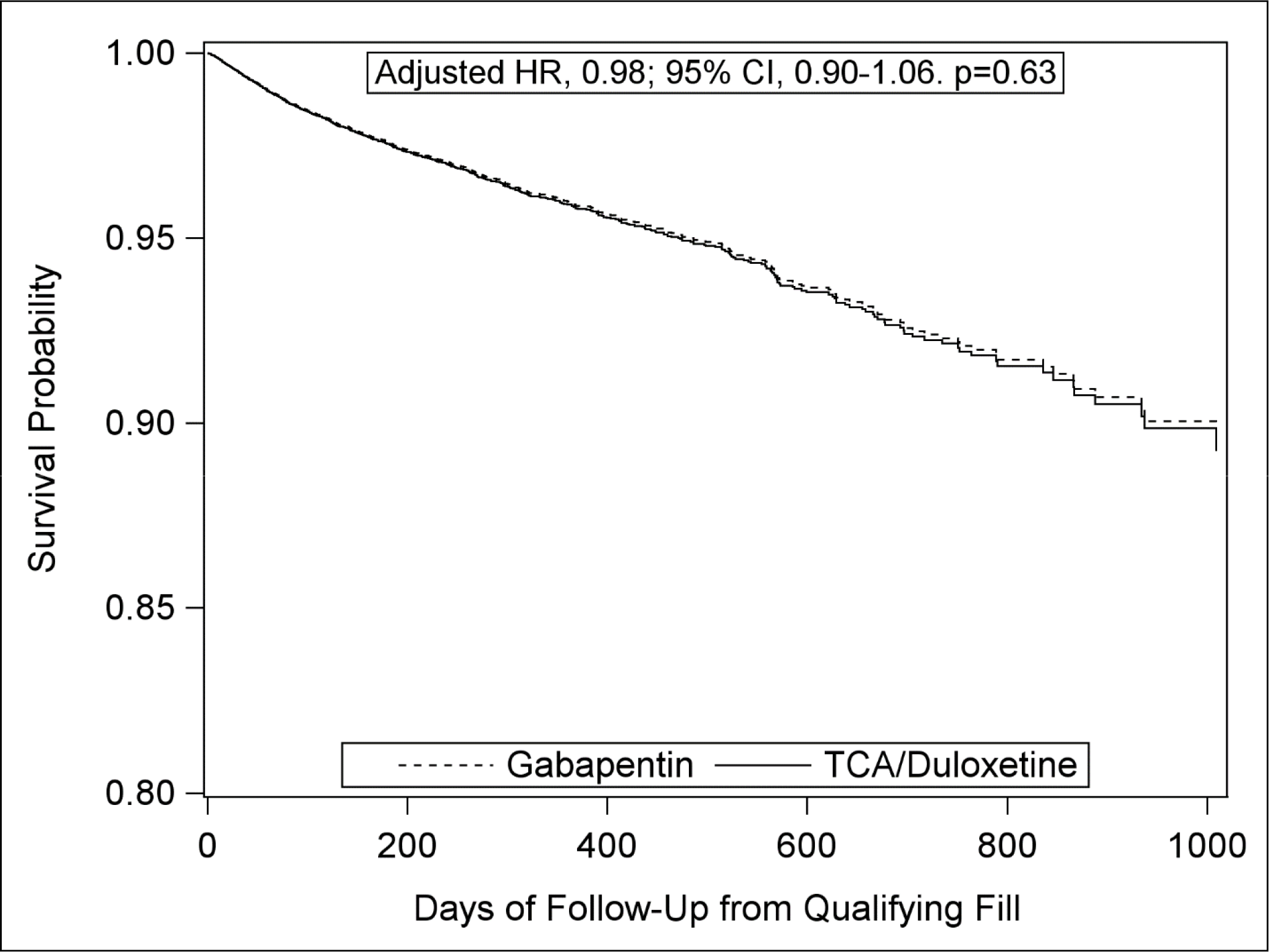
Primary analysis Cox proportional hazard curves for any-time mortality.

Results of the secondary analysis requiring ≥2 fills of each medication of interest were similar to those of the primary analysis. A total of 165,585 people who filled ≥2 gabapentin and opioid prescriptions were eligible to be matched with 42,344 people who filled ≥2 TCA/duloxetine and opioid prescriptions; of these, 15,593 were in each matched group (eFigure 10). The groups were well-matched (eTable 8). We observed similar proportions of death and slightly greater proportions of medical complications among people treated with gabapentin at any time (eTable 9). We did not observe any significant difference in risk of mortality overall (eFigures 11-12; aHR, 1.01; 95% CI, 0.86-1.20) or when stratified by average MME category (eTable 10). There was no significant increased risk of any-time major medical complications (eFigures 13-14; aHR, 1.04; 95% CI, 0.99-1.08). Results for 30-day outcomes were similar (eTable 11; eFigures 15-18).

## Discussion

Among Medicare participants with a spine-related diagnosis between 2017 and 2019, we found no difference in mortality risk among people who concurrently filled prescriptions for gabapentin + opioids versus an active control + opioids. This finding was consistent even when we restricted our sample to people who filled ≥2 prescriptions for gabapentin/active control + opioids and therefore were more likely to have consumed their medications. We did, however, detect a small but statistically significantly greater risk of a major medical complication at any time among those who filled gabapentin + opioids compared to those who filled TCA/duloxetine + opioids in our primary analysis.

A recent study by Corriere et al. used inverse probability of treatment weighting (ITPW) and an active control design to compare Medicare beneficiaries who filled prescriptions for gabapentin to those who filled duloxetine from 2015-2018.^10^ Unlike the current study in which there was no increased mortality risk overall or in any subgroup, Corriere et al. found twice the mortality risk with gabapentin vs. duloxetine (aHR, 2.0; 95% CI, 1.2-3.5) among people with the highest opioid doses (≥50 MME/day), but no increased risk among people with no or low-dose opioids (<50 MME/day). This difference may be explained by methodological differences between these studies. We used the 100% Medicare population of people with spine-related diagnoses and required all participants to have concurrent opioid prescriptions. In contrast, Corriere et al. analyzed a 20% Medicare sample of people with non-cancer chronic pain, in which approximately half of the sample did <u>not</u> use opioids concurrently with gabapentin. We matched on opioid dose using 9 MME categories, in contrast to the 3 MME categories used by Corriere et al., which might have resulted in residual confounding. We also matched exactly on two other factors to ensure comparability between groups: the duration of time between opioid and concurrent medication (gabapentin or TCA/duloxetine) fills and major medical complications immediately before the start of follow-up. In extensive *post hoc* analyses we conducted to emulate the methods of Corriere et al., we were unable to replicate their finding of elevated mortality risk in people with the highest MME. However, a sensitivity analysis by Corriere et al. using PSM produced findings similar to ours, showing no effect of gabapentin on all-cause mortality (aHR 1.3; 95% CI 0.6-5.4). This suggests that the differences between study findings may be explained by the analytic methods used by Corriere et al. (IPTW and risk sets determined based on time since initiating each MME category) vs. the current study (PSM and risk sets determined based on time since initiating gabapentin/active control). While the strengths and weaknesses of IPTW vs. PSM are actively debated, IPTW can produce imprecise treatment effect estimates with undue influence of certain observations in some situations,^32,33^ which may explain the different results.

While other studies have found that people exposed to gabapentin were at increased risk of mortality compared to people not exposed to gabapentin, it is difficult to draw conclusions from these due to their limited control for potential confounding. A case-control study of opioid users in Ontario, Canada who died of opioid-related causes from 1997-2013 found a 50% greater adjusted odds of mortality associated with gabapentin prescriptions compared to those without in the 4 months prior to death.^7^ Though that study adjusted for opioid dose and a few comorbid conditions in the 3 years prior to death, the analysis did not include an active control group, potentially leading to confounding by indication if people who filled prescriptions for gabapentin + opioids were at greater risk of mortality compared to people who only filled prescriptions for opioids without the need for a second medication. Furthermore, a recent study^8^ examining mortality among dialysis patients found no difference in mortality risk among patients with prescriptions for an opioid and gabapentin compared to those prescribed only opioids, consistent with our results.

The clinical implication of our findings is that the risk of death among older adults with spine-related conditions who fill opioids is not increased by adding gabapentin compared to adding an active comparator medication. However, the finding that adding gabapentin to opioids is associated with a small yet significantly increased risk of medical complications warrants further study, particularly in light of other recent studies which found associations between gabapentin prescriptions and pulmonary complications.^21,34,35^ Because medical complications was our secondary outcome, these results must be interpreted cautiously.

A major strength of our study is that we pre-specified a detailed analysis plan^12^ prior to analyzing this large, representative cohort of older US adults and created comparison groups that were alike in many ways. We also used an active control group of patients using TCAs/duloxetine and opioids concurrently to minimize confounding by the need for an additional analgesic and to mirror the clinical situation that, once a decision has been made that opioids alone are insufficient, a relevant choice is whether the additional analgesic is gabapentin, or another analgesic. However, it is possible that residual confounding remained. It is possible, for example, that if we analyzed only people who were exposed to particularly high dosages of gabapentin or TCA/duloxetine plus opioids, we may have found different results. Because our cohort included only older adults with spine-related conditions, including back and neck pain, we cannot be certain that our results would apply to younger populations or people with different pain conditions.

We found no increased mortality risk among people who filled gabapentin + opioids compared to those who filled an active control + opioids, which should reassure providers seeking options for pain control, particularly patients with spine diagnoses. However, a small but significantly increased risk of medical complications among those who filled gabapentin + opioids compared to active controls + opioids suggests that patients should be closely monitored while they are on these medications concurrently.

## Supporting information

ETable 3

## Data Availability

All data produced in the present work are contained in the manuscript.

