## Supplemental tables and figures for "Mortality with concurrent treatment with gabapentin and opioids among people with spine diagnoses in the U.S. Medicare population: a propensity-matched cohort study"

**eTable 1**. Codes included in the study cohort and indication of whether the code is associated with neuropathic pain.^a^

| ICD-10 Code | Code Description | Neuropathic Pain (0=no; 1=yes) |
| --- | --- | --- |
| G54.0 | Brachial plexus disorder | 1 |
| G54.1 | Lumbosacral plexus disorder | 1 |
| G54.2 | Cervical root disorders | 1 |
| G54.3 | Thoracic root disorders | 1 |
| G54.4 | Lumbosacral root disorders | 1 |
| M40.X | Kyphosis and lordosis | 0 |
| M41.X | Scoliosis | 0 |
| M42.X | Spinal osteochrondrosis | 0 |
| M43.0X | Other deforming dorsopathies | 0 |
| M43.1X | Spondylolisthesis | 1 |
| M43.2X | Fusion of spine | 0 |
| M43.4 | Other recurrent atlantoaxial dislocation | 0 |
| M43.5X | Other recurrent vertebral dislocation | 0 |
| M43.6 | Torticollis | 0 |
| M43.8X | Other specified deforming dorsopathies | 0 |
| M43.9 | Deforming dorsopathy, unspecified | 0 |
| M47.2X | Other spondylosis with radiculopathy | 1 |
| M47.81X | Spondylosis without myelopathy or radiculopathy | 0 |
| M47.89X | Other spondylosis | 0 |
| M47.9 | Spondylosis, unspecified | 0 |
| M48.00 | Spinal stenosis, site unspecified | 1 |
| M48.01 | Spinal stenosis, occipito-atlanto-axial region | 1 |
| M48.02 | Spinal stenosis, cervical region | 1 |
| M48.03 | Spinal stenosis, cervico-thoracic region | 1 |
| M48.04 | Spinal stenosis, thoracic region | 1 |
| M48.05 | Spinal stenosis, thoracolumbar region | 1 |
| M48.061 | Spinal stenosis, lumbar region, without neurogenic claudication | 1 |
| M48.062 | Spinal stenosis, lumbar region, with neurogenic claudication | 1 |
| M48.1X | Ankylosing hyperostosis | 0 |
| M48.2X | Kissing spine | 0 |
| M48.8X | Other specified spondylopathies | 0 |
| M48.9 | Spondylopathy, unspecified | 0 |
| M49.8X | Spondylopathy in diseases classified elsewhere | 0 |
| M50.1X | Cervical disc disorder with radiculopathy | 1 |
| M50.2X | Other cervical disc displacement | 0 |
| M50.3X | Other cervical disc degeneration | 0 |
| M50.8X | Other cervical disc disorders | 0 |
| M50.9X | Cervical disc disorder, unspecified | 0 |
| M51.1X | Thoracic, thoracolumbar and lumbosacral intervertebral disc disorders with radiculopathy | 1 |
| M51.2X | Other thoracic, thoracolumbar and lumbosacral intervertebral disc displacement | 0 |
| M51.3X | Other thoracic, thoracolumbar and lumbosacral intervertebral disc degeneration | 0 |
| M51.4X | Schmorl's nodes | 0 |
| M51.8X | Other thoracic, thoracolumbar and lumbosacral intervertebral disc disorders | 0 |
| M51.9 | Unspecified thoracic, thoracolumbar and lumbosacral intervertebral disc disorder | 0 |
| M51.AX | Other lumbar and lumbosacral annulus fibrosus disc defects | 0 |
| M53.0 | Cervicocranial syndrome | 0 |
| M53.1 | Cervicobrachial syndrome | 1 |
| M53.2X | Spinal instabilities | 0 |
| M53.3 | Sacrococcygeal disorders, not elsewhere classified | 0 |
| M53.8X | Other specified dorsopathies | 0 |
| M53.9 | Dorsopathy, unspecified | 0 |
| M54.0X | Panniculitis affecting regions of the neck and back | 0 |
| M54.1X | Radiculopathy | 1 |
| M54.2 | Cervicalgia | 0 |
| M54.3X | Sciatica | 1 |
| M54.4X | Lumbago with sciatica | 1 |
| M54.5X | Low back pain | 0 |
| M54.6 | Pain in thoracic spine | 0 |
| M54.8X | Other dorsalgia | 1 |
| M54.9 | Dorsalgia, unspecified | 0 |
| S13.4X | Sprains of ligaments of cervical spine | 0 |
| S13.5X | Sprain of thyroid region | 0 |
| S13.8X | Sprain of joints and ligaments of other parts of neck | 0 |
| S13.9X | Sprain of joints and ligaments of unspecified parts of neck | 0 |
| S14.2X | Injury of nerve root of cervical spine | 1 |
| S14.3X | Injury of brachial plexus | 1 |
| S14.4X | Injury of peripheral nerves of neck | 1 |
| S14.5X | Injury of cervical sympathetic nerves | 1 |
| S14.8X | Injury of other specified nerves of neck | 1 |
| S14.9X | Injury of other unspecified nerves of neck | 1 |
| S19.8X | Other specified injuries of the neck | 0 |
| S22.2X | Fracture of sternum | 0 |
| S22.3X | Fracture of 1 rib | 0 |
| S22.4X | Fracture of multiple ribs | 0 |
| S22.5X | Flail chest | 0 |
| S22.9X | Fracture of bony thorax, part unspecified | 0 |
| S23.3X | Sprain of ligaments of thoracic spine | 0 |
| S23.4X | Sprain of ribs and sternum | 0 |
| S23.8X | Sprain of other specified parts of thorax | 0 |
| S23.9X | Sprain of unspecified parts of thorax | 0 |
| S24.2X | Injury of nerve root of thoracic spine | 1 |
| S24.3X | Injury of peripheral nerves of thorax | 1 |
| S24.4X | Injury of thoracic sympathetic nervous system | 1 |
| S24.8X | Injury of other specified nerves of thorax | 1 |
| S24.9X | Injury of other unspecified nerves of thorax | 1 |
| S29.00X | Unspecified injury of muscle and tendon at thorax level | 0 |
| S29.01X | Strain of muscle and tendon of thorax | 0 |
| S29.02X | Laceration of muscle and tendon of thorax | 0 |
| S29.09X | Other injury of muscle and tendon of thorax | 0 |
| S29.8X | Other specified injuries of thorax | 0 |
| S29.9X | Unspecified injury of thorax | 0 |
| S33.5X | Sprain of ligaments of lumbar spine | 0 |
| S33.6X | Sprain of sacroiliac joint | 0 |
| S33.8X | Sprain of other parts of lumbar spine and pelvis | 0 |
| S33.9X | Sprain of unspecified parts of lumbar spine and pelvis | 0 |
| S34.2X | Injury of nerve root of lumbar and sacral spine | 1 |
| S34.4X | Injury of lumbosacral plexus | 1 |
| S34.5X | Injury of lumbar, sacral, and pelvic sympathetic nerves | 1 |
| S34.6X | Injury of peripheral nerves at abdomen, lower back, and pelvis level | 1 |
| S34.8X | Injury of other specified nerves at abdomen, lower back, and pelvis level | 1 |
| S34.9X | Injury of other unspecified nerves at abdomen, lower back and pelvis level | 1 |
| S39.002X | Unspecified injury of muscle, fascia and tendon of lower back | 0 |
| S39.012X | Strain of muscle, fascia and tendon of lower back | 0 |
| S39.022X | Laceration of muscle, fascia and tendon of lower back | 0 |
| S39.092X | Other injury of muscle, fascia and tendon of lower back | 0 |
| S39.82X | Other specified injuries of lower back | 0 |
| S39.92X | Unspecified injury of lower back | 0 |

^a^Spine-related neuropathic pain was a covariate as it was expected to be more prevalent in those receiving gabapentin.

eTable 2. People with these codes were *excluded* if these diagnoses occurred before index fill date.

| ICD-10 Code | Code Description |
| --- | --- |
| C41.2 | Malignant neoplasm of vertebral column |
| C72.0 | Malignant neoplasm of spinal cord |
| C72.1 | Malignant neoplasm of cauda equina |
| D43.4 | Neoplasm of uncertain behavior of spinal cord |
| G06.1 | Instraspinal abscess and granuloma |
| G40.X | Epilepsy and recurrent seizures |
| M43.3 | Recurrent atlantoaxial dislocation with myelopathy |
| M46.2X | Osteomyelitis of vertebra |
| M46.3X | Infection of intervertebral disc |
| M46.4X | Discitis, unspecified |
| M46.5X | Other infective spondylopathies |
| M47.01X | Anterior spinal artery compression syndromes |
| M47.02X | Vertebral artery compression syndromes |
| M47.1X | Other spondylosis with myelopathy |
| M48.3X | Traumatic spondylopathy |
| M48.4X | Fatigue fracture of vertebra |
| M48.5X | Collapsed vertebra, not elsewhere classified |
| M50.0X | Cervical disc disorder with myelopathy |
| M51.0X | Thoracic, thoracolumbar and lumbosacral intervertebral disc disorders with myelopathy |
| R56.X | Convulsions, not elsewhere classified |
| S12.X | Fracture of cervical vertebra and other parts of neck |
| S13.0X | Traumatic rupture of cervical intervertebral disc |
| S13.1X | Subluxation and dislocation of cervical vertebrae |
| S13.2X | Dislocation of other and unspecified parts of neck |
| S14.0X | Concussion and edema of cervical spinal cord |
| S14.10X | Unspecified injuries of cervical spinal cord |
| S14.11X | Complete lesion of cervical spinal cord |
| S14.12X | Central cord syndrome of cervical spinal cord |
| S14.13X | Anterior cord syndrome of cervical spinal cord |
| S14.14X | Brown-Sequard syndrome of cervical spinal cord |
| S14.15X | Other incomplete lesions of cervical spinal cord |
| S22.0X | Fracture of thoracic vertebra |
| S23.0X | Traumatic rupture of thoracic intervertebral disc |
| S23.1X | Subluxation and dislocation of thoracic vertebra |
| S23.2X | Dislocation of other and unspecified parts of thorax |
| S24.0X | Concussion and edema of thoracic spinal cord |
| S24.10X | Unspecified injuries of thoracic spinal cord |
| S24.11X | Complete lesion of thoracic spinal cord |
| S24.13X | Anterior cord syndrome of thoracic spinal cord |
| S24.14X | Brown-Sequard syndrome of thoracic spinal cord |
| S24.15X | Other incomplete lesions of thoracic spinal cord |
| S32.0X | Fracture of lumbar vertebra |
| S32.1X | Fracture of sacrum |
| S32.2X | Fracture of coccyx |
| S32.3X | Fracture of ileum |
| S32.4X | Fracture of acetabulum |
| S32.5X | Fracture of pubis |
| S32.6X | Fracture of ischium |
| S32.8X | Fracture of other parts of the pelvis |
| S32.9X | Fracture of unspecified parts of lumbosacral spine and pelvis |
| S33.0X | Traumatic rupture of lumbar intervertebral disc |
| S33.1X | Subluxation and dislocation of lumbar vertebra |
| S33.2X | Dislocation of sacroiliac and sacrococcygeal joint |
| S33.3X | Dislocation of other and unspecified parts of lumbar spine and pelvis |
| S33.4X | Traumatic rupture of symphysis pubis |
| S34.0X | Concussion and edema of lumbar and sacral spinal cord |
| S34.10X | Unspecified injuries of lumbar spinal cord |
| S34.11X | Complete lesion of lumbar spinal cord |
| S34.12X | Incomplete lesion of lumbar spinal cord |
| S34.13X | Other and unspecified injury to sacral spinal cord |
| S34.3X | Injury of cauda equina |
| T84.63X | Infection and inflammatory reaction due to internal fixation device |

eTable 3 is provided separately as a supplemental document.

eFigures 1a-1d. Timelines of washout periods, index dates, qualifying dates, and follow-up periods.

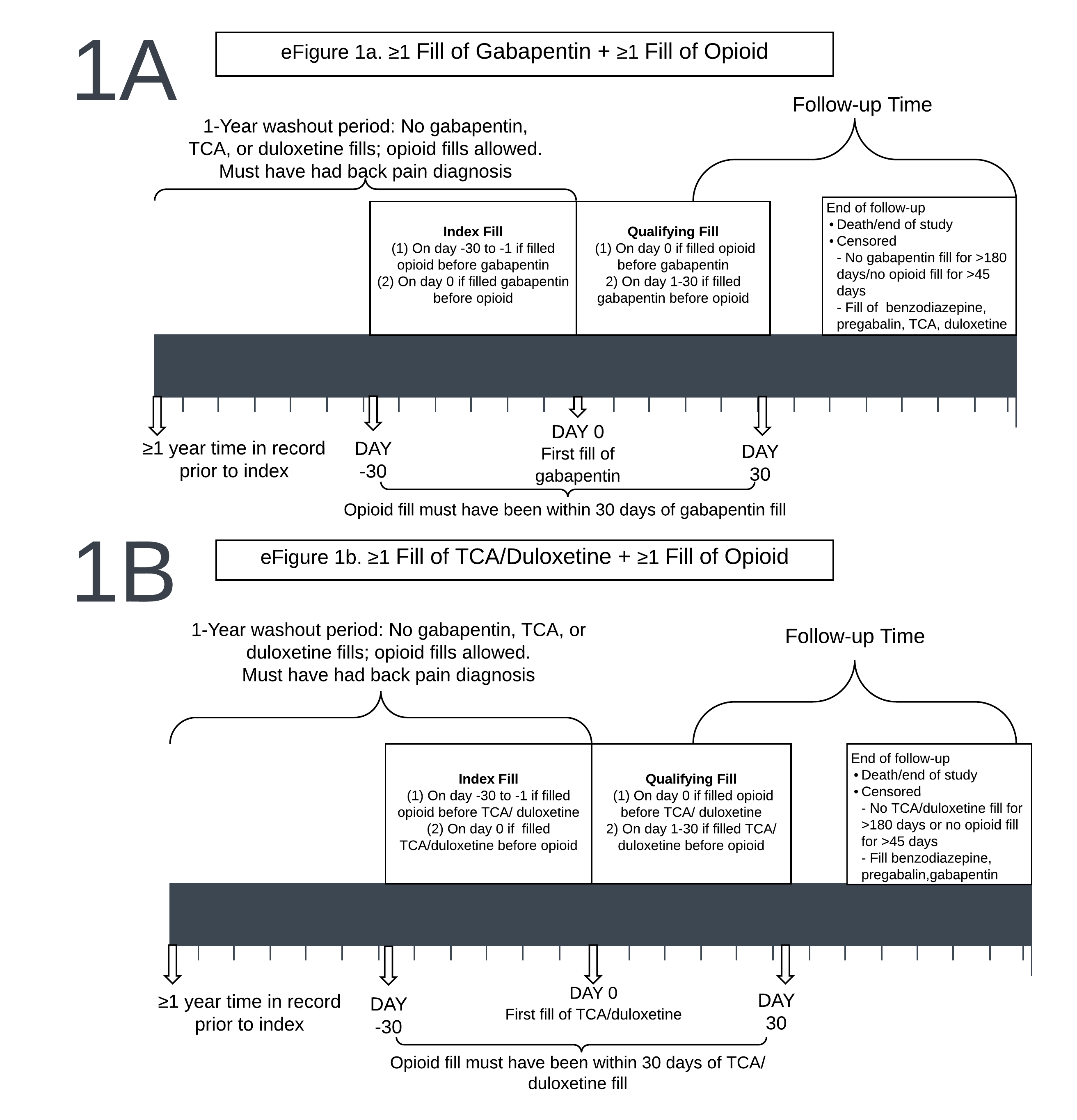

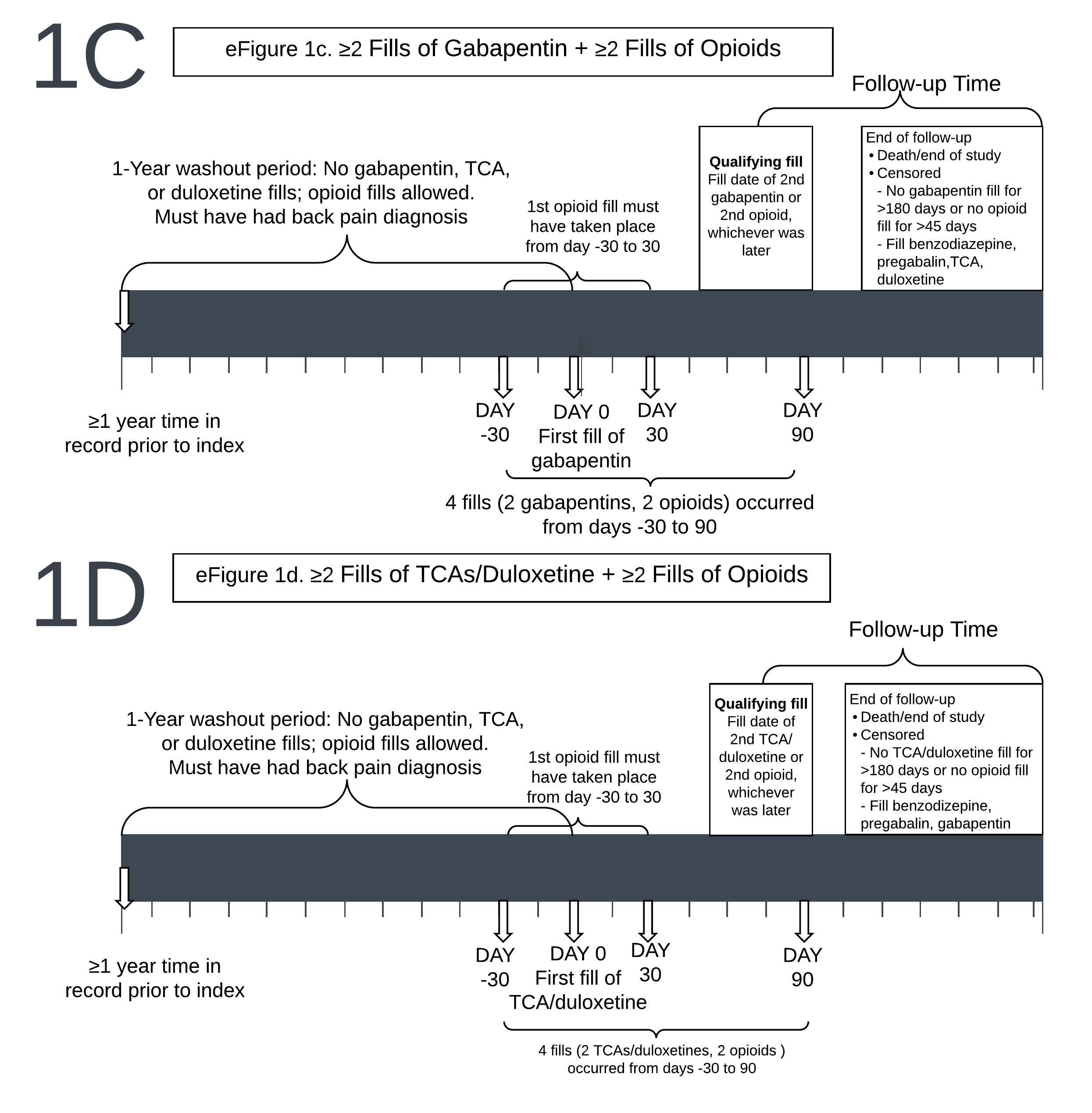
Table e4. List of major medical complications by category. Levels of acute risk were determined by model weights from earlier research examining mortality in Medicare patients with osteoporotic vertebral fractures and from expert clinical opinions of co-authors (JLF; RAD; MC; KW; JGJ; PS).

- *High acute risk*- any complication listed in this subgroup could be matched with another complication in the same subgroup:
  - Cardiac arrest (ICD-10 diagnosis codes I46.2, I46.8, I46.9, I97.88, I97.89)
  - Intubation/mechanical ventilation/CPR (ICD-10 procedure codes 0BH17EZ, 0BH18EZ, 5A1935Z, 5A1945Z, 5A1955Z, 5A12012
  - Respiratory failure (ICD-10 diagnosis codes J95.0X, J95.2, J96.90, J80, R06.03)
- *Moderate acute risk-* any complication listed in this subgroup could be matched with another complication in the same subgroup:
  - Sepsis (ICD-10 diagnosis codes A02.1, A26.7, A32.7, A40.X, A41.X, A42.7, R65.X, T81.44X)
  - Pneumonia (ICD-10 diagnosis codes J12.9, J13, J14, J15.X, J18.X, J69.X)
  - Fluid and electrolyte disorders (ICD-10 diagnosis codes E22.2, E86.X, E87.X)
  - Weight loss (ICD-10 diagnosis codes E40.X-E46.X, E64.0, R63.4, R64)
  - Cancer (ICD-10 diagnosis codes C00.X -C26.X, C30.X -C34.X, C37.X -C41.X, C43.X, C45.X-C58.X, C60.X-C75.X, C81.X-C94.3X, C94.8X, C95.X, C96.0X-C96.4X, C96.9, C96.A, C96.Z, D45, D89, Z8546)
  - Acute myocardial infarction (ICD-10 diagnosis codes I.21X)
  - Paralysis (ICD-10 diagnosis codes G04.1, G11.4, G80.1, G80.2, G81.X-G83.4X, G83.9)
- *Low acute risk*- any complication listed in this subgroup could be matched with another complication in the same subgroup:
  - Coagulopathy (ICD-10 diagnosis codes D65.X-D68.X, D69.1, D69.3-D69.6)
  - Gastrointestinal bleed (ICD-10 diagnosis codes K29.01, K29.21, K29.31, K29.41, K29.51, K29.61, K29.71, K29.81, K29.91, K92.2,)
  - Cardiac arrhythmias (ICD-10 diagnosis codes I44.0, I44.1, I44.3X-I45.2X, I45.4X-I45.8X, I45.9, I47X-I49X, R00.0, R00.1, R00.8, Z95.0, Z95.810, Z95.818, Z95.9)
  - Stroke (ICD-10 diagnosis codes G46, G46.0, G46.1, G46.2, G46.3, G46.4, G46.5, G46.6, G46.7, G97.8X, I60.X, I61.X, I63.X, I64.X)

eFigure 2. Replicating selected inclusion and exclusion criteria used in the cohort study by Corriere et al. for *post hoc* analysis.

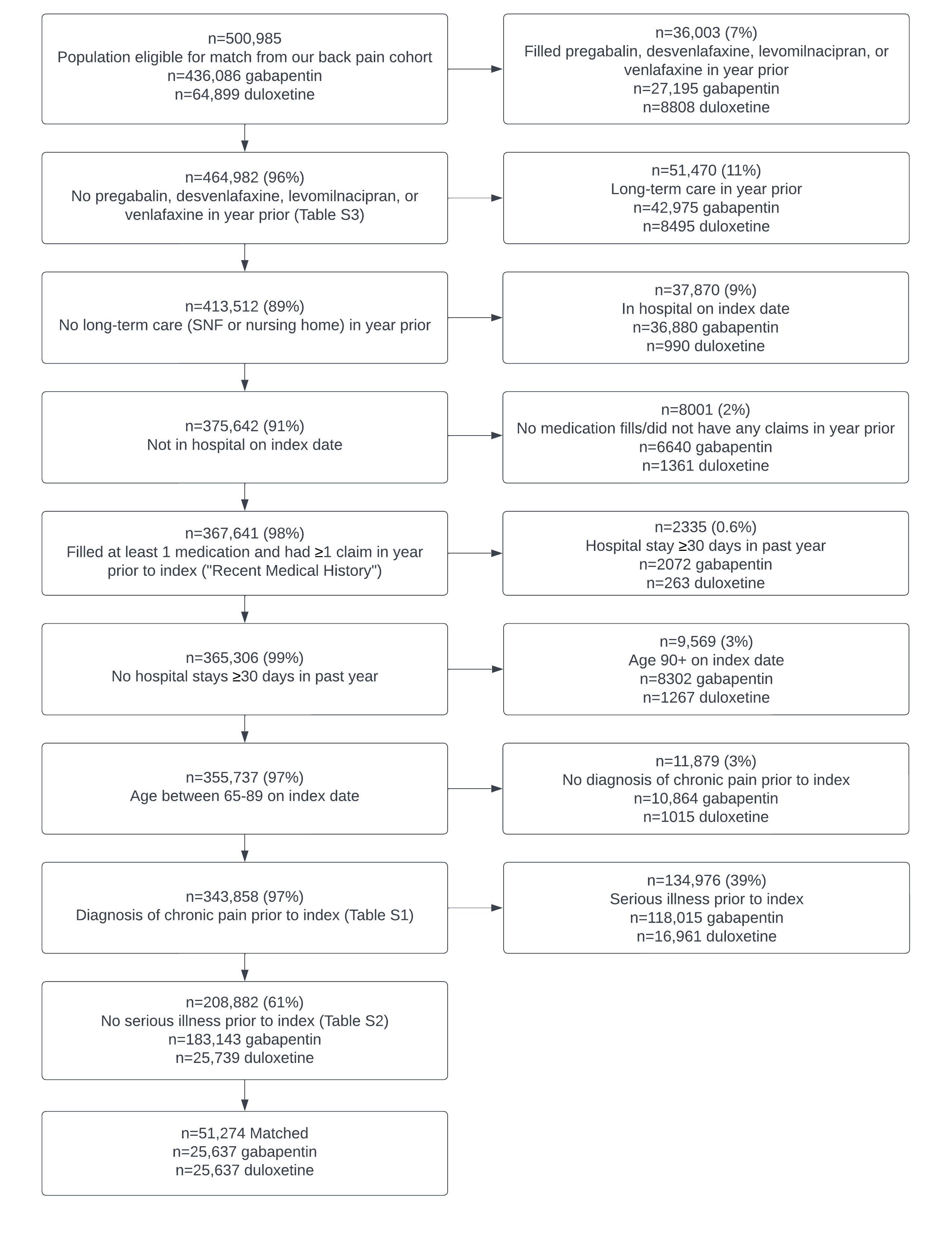

eTable 5. Standardized differences comparing patients who received gabapentin to those who received TCAs/duloxetine in the unmatched and matched populations for the primary analysis.

|  | **Primary Analysis Unmatched Population** | | | **Primary Analysis Matched Population** | | |
| --- | --- | --- | --- | --- | --- | --- |
|  | **Gabapentin n=426,216 (81%) n (%)** | **TCA/Duloxetine n=99,668 (19%)**  **n (%)** | **Standardized difference** | **Gabapentin n=67,133 (50%) n (%)** | **TCA/Duloxetine n=67,133 (50%)**  **n (%)** | **Standardized difference** |
| ***DEMOGRAPHIC VARIABLES*** |  |  |  |  |  |  |
| *Age in years (median, IQR)* | 73.7 (69.5, 79.3) | 73.2 (69.0, 79.2) | -0.03 | 73.5 (69.4, 79.2) | 73.2 (69.0, 79.2) | -0.02 |
| *Female* | 259,848 (61.0) | 68,020 (68.2) | -0.15 | 45,413 (67.6) | 44,141 (65.8) | 0.04 |
| *Patient race* |  |  |  |  |  |  |
| *Unknown* | 6,231 (1.5) | 998 (1.0) | -0.04 | 703 (1.0) | 785 (1.2) | 0.01 |
| *White* | 365,163 (85.7) | 87,609 (87.9) | 0.07 | 58,655 (87.4) | 58,340 (86.9) | -0.01 |
| *Black* | 34,970 (8.2) | 7,360 (7.4) | -0.03 | 5,102 (7.6) | 5,282 (7.9) | 0.01 |
| *Other* | 4,538 (1.1) | 843 (0.8) | -0.02 | 569 (0.8) | 607 (0.9) | 0.01 |
| *Asian* | 6,036 (1.4) | 830 (0.8) | -0.06 | 645 (1.0) | 658 (1.0) | 0.00 |
| *Hispanic* | 7,180 (1.7) | 1,388 (1.4) | -0.02 | 1,026 (1.5) | 1,058 (1.6) | 0.00 |
| *North American Native* | 2,098 (0.5) | 640 (0.6) | 0.02 | 433 (0.6) | 403 (0.6) | -0.01 |
| *Region* |  |  |  |  |  |  |
| *New England* | 19,931 (4.7) | 3,717 (3.7) | -0.05 | 2,572 (3.8) | 2,710 (4.0) | 0.01 |
| *NY/NJ/PR* | 30,749 (7.2) | 5,994 (6.0) | -0.05 | 4,082 (6.1) | 4,227 (6.3) | 0.01 |
| *PA/DE/MD/DC/WV/VA* | 40,549 (9.5) | 9,125 (9.2) | -0.01 | 6,107 (9.1) | 6,239 (9.3) | 0.01 |
| *KY/TN/NC/SC/GA/AL/MS/FL* | 101,462 (23.8) | 26,001 (26.1) | 0.05 | 17,216 (25.6) | 17,058 (25.4) | -0.01 |
| *MN/WI/MI/OH/IN/IL* | 69,797 (16.4) | 16,558 (16.6) | 0.01 | 11,092 (16.5) | 10,983 (16.4) | 0.00 |
| *NM/TX/OK/AR/LA* | 57,179 (13.4) | 13,204 (13.2) | 0.00 | 9,017 (13.4) | 9,056 (13.5) | 0.00 |
| *NE/IA/KS/MO* | 25,127 (5.9) | 6,150 (6.2) | 0.01 | 4,068 (6.1) | 4,079 (6.1) | 0.00 |
| *MT/ND/SD/WY/UT/CO* | 12,982 (3.0) | 3,334 (3.3) | 0.02 | 2,226 (3.3) | 2,199 (3.3) | 0.00 |
| *CA/HI/NV/AZ/Pac Islands* | 52,126 (12.2) | 11,316 (11.4) | -0.03 | 7,911 (11.8) | 7,855 (11.7) | 0.00 |
| *AK/WA/OR/ID* | 16,311 (3.8) | 4,268 (4.3) | 0.02 | 2,842 (4.2) | 2,727 (4.1) | -0.01 |
| *Buy-in from state* | 82,043 (19.2) | 23,872 (24.0) | 0.11 | 15,191 (22.6) | 15,166 (22.6) | 0.00 |
| *Index^a^ year* |  |  |  |  |  |  |
| *2017* | 162,504 (38.1) | 41,374 (41.5) | 0.07 | 27,673 (41.2) | 27,390 (40.8) | -0.01 |
| *2018* | 141,075 (33.1) | 32,051 (32.2) | -0.02 | 21,785 (32.5) | 21,719 (32.4) | 0.00 |
| *2019* | 122,637 (28.8) | 26,243 (26.3) | -0.05 | 17,675 (26.3) | 18,024 (26.8) | 0.01 |
| ***CLINICAL VARIABLES*** |  |  |  |  |  |  |
| *Days from first back pain diagnosis to index* | 473 (212, 778) | 532 (331, 824) | 0.17 | 528 (304, 820) | 511 (304, 800) | -0.02 |
| Days from most recent back pain diagnosis to index | 12 (0, 141) | 31 (1, 172) | 0.05 | 15 (0, 160) | 29 (1, 166) | -0.01 |
| *Charlson co-morbidity score (median, IQR)* | 2 (1, 4) | 2 (1, 4) | -0.01 | 2 (1, 4) | 2 (1, 4) | 0.02 |
| *0* | 96,144 (22.6) | 21,344 (21.4) | -0.03 | 15,131 (22.5) | 15,064 (22.4) | 0.00 |
| *1-2* | 147,048 (34.5) | 34,924 (35.0) | 0.01 | 24,059 (35.8) | 23,609 (35.2) | -0.01 |
| *3-4* | 88,684 (20.8) | 21,742 (21.8) | 0.01 | 14,226 (21.2) | 14,345 (21.4) | -0.01 |
| *5+* | 94,340 (22.1) | 21,658 (21.7) | -0.01 | 13,717 (20.4) | 14,115 (21.0) | 0.01 |
| *Diagnoses/procedures (yes/no) in year prior to index* |  |  |  |  |  |  |
| *Cancer (excluding skin cancer)* | 80,680 (18.9) | 15,565 (15.6) | -0.09 | 10,124 (15.1) | 10,530 (15.7) | 0.02 |
| *Metastatic cancer* | 19,281 (4.5) | 3,258 (3.3) | -0.06 | 2,024 (3.0) | 2,239 (3.3) | 0.02 |
| *Congestive heart failure* | 87,268 (20.5) | 21,416 (21.5) | 0.02 | 13,640 (20.3) | 13,686 (20.4) | 0.00 |
| *Dementia* | 20,494 (4.8) | 7,073 (7.1) | 0.10 | 4,091 (6.1) | 4,086 (6.1) | 0.00 |
| *Weight loss* | 25,501 (6.0) | 7,637 (7.7) | 0.07 | 4,503 (6.7) | 4,403 (6.6) | -0.01 |
| *Hemiplegia* | 6,184 (1.5) | 1,566 (1.6) | 0.01 | 942 (1.4) | 977 (1.5) | 0.00 |
| *Alcohol abuse* | 2,197 (0.5) | 580 (0.6) | 0.01 | 344 (0.5) | 370 (0.6) | 0.01 |
| *Cardiac arrhythmias* | 138,822 (32.6) | 31,837 (31.9) | -0.01 | 20,350 (30.3) | 20,583 (30.7) | 0.01 |
| *Chronic pulmonary disease* | 128,094 (30.1) | 33,301 (33.4) | 0.07 | 21,465 (32.0) | 21,368 (31.8) | 0.00 |
| *Coagulopathy* | 22,553 (5.3) | 4,916 (4.9) | -0.02 | 3,121 (4.6) | 3,166 (4.7) | 0.00 |
| *Diabetes with complications* | 93,342 (21.9) | 21,102 (21.2) | -0.02 | 14,110 (21.0) | 14,423 (21.5) | 0.01 |
| *Deficiency anemia* | 99,764 (23.4) | 25,235 (25.3) | 0.04 | 16,257 (24.2) | 16,193 (24.1) | 0.00 |
| *Fluid/electrolyte disorders* | 73,572 (17.3) | 18,794 (18.9) | 0.04 | 11,679 (17.4) | 11,713 (17.4) | 0.00 |
| *Liver disease* | 32,114 (7.5) | 7,835 (7.9) | 0.01 | 5,107 (7.6) | 5,149 (7.7) | 0.00 |
| *Peripheral vascular disease* | 98,553 (23.1) | 23,313 (23.4) | 0.01 | 15,217 (22.7) | 15,529 (23.1) | 0.01 |
| *Pulmonary circulation disorders* | 23,319 (5.5) | 5,829 (5.8) | 0.02 | 3,753 (5.6) | 3,665 (5.5) | -0.01 |
| *HIV/AIDS* | 752 (0.2) | 187 (0.2) | 0.00 | 132 (0.2) | 118 (0.2) | 0.00 |
| *Hypertension* | 336,887 (79.0) | 78,398 (78.7) | -0.01 | 52,852 (78.7) | 52,625 (78.4) | -0.01 |
| *Myocardial infarction* | 12,288 (2.9) | 2,906 (2.9) | 0.00 | 1,853 (2.8) | 1,858 (2.8) | 0.00 |
| *Rheumatic disease* | 28,994 (6.8) | 9,627 (9.7) | 0.10 | 5,810 (8.7) | 5,702 (8.5) | -0.01 |
| *Peptic ulcer disease* | 9,880 (2.3) | 2,736 (2.7) | 0.03 | 1,743 (2.6) | 1,721 (2.6) | 0.00 |
| *Diabetes without complications* | 141,015 (33.1) | 31,539 (31.6) | -0.03 | 21,274 (31.7) | 21,651 (32.3) | 0.01 |
| *Opioid use disorder* | 52,513 (12.3) | 22,359 (22.4) | 0.27 | 12,634 (18.8) | 13,093 (19.5) | 0.02 |
| *Parkinson’s disease* | 6,305 (1.5) | 1,733 (1.7) | 0.02 | 1,190 (1.8) | 1,104 (1.6) | -0.01 |
| *Obesity* | 77,772 (18.2) | 17,959 (18.0) | -0.01 | 11,719 (17.5) | 12,085 (18.0) | 0.01 |
| *Insomnia* | 42,318 (9.9) | 15,220 (15.3) | 0.16 | 8,880 (13.2) | 8,714 (13.0) | -0.01 |
| *Sleep apnea* | 58,310 (13.7) | 14,487 (14.5) | 0.02 | 9,351 (13.9) | 9,403 (14.0) | 0.00 |
| *Renal failure* | 79,987 (18.8) | 18,985 (19.0) | 0.01 | 12,477 (18.6) | 12,459 (18.6) | 0.00 |
| *Electroconvulsive therapy* | 51 (0.0) | 43 (0.0) | 0.02 | 14 (0.0) | 15 (0.0) | 0.00 |
| *Therapeutic repetitive transcranial magnetic stimulation treatment* | 65 (0.0) | 34 (0.0) | 0.01 | 14 (0.0) | 19 (0.0) | 0.00 |
| *Spine-related neuropathic pain* (see eTable 1) | 244,593 (57.4) | 48,973 (49.1) | -0.17 | 34,146 (50.9) | 35,338 (52.6) | 0.04 |
| *Number of inpatient admissions (median, IQR)* | 0 (0, 1) | 0 (0, 1) | -0.09 | 0 (0, 1) | 0 (0, 1) | 0.03 |
| *Number of outpatient encounters (median, IQR)* | 24 (15, 38) | 25 (15, 39) | -0.06 | 24 (15, 38) | 24 (14, 38) | 0.01 |
| *Number of unique dates with generalized anxiety disorder diagnoses (median, IQR)* | 0 (0, 0) | 0 (0, 0) | 0.02 | 0 (0, 0) | 0 (0, 0) | 0.01 |
| *Number of unique dates with post-traumatic stress disorder diagnoses (median, IQR)* | 0 (0, 0) | 0 (0, 0) | 0.10 | 0 (0, 0) | 0 (0, 0) | -0.01 |
| *Number of unique dates with back pain diagnoses (median, IQR)* | 3 (1, 8) | 4 (1, 9) | 0.07 | 4 (1, 9) | 4 (1, 9) | 0.01 |
| *Number of unique dates with non-spine neuropathy diagnoses (median, IQR)* | 0 (0, 0) | 0 (0, 0) | 0.01 | 0 (0, 0) | 0 (0, 0) | 0.01 |
| *Number of unique dates with depression diagnoses (median, IQR)* | 0 (0, 0) | 0 (0, 1) | 0.01 | 0 (0, 0) | 0 (0, 1) | 0.01 |
| *Medication fills in year prior to index* |  |  |  |  |  |  |
| *NSAID* | 161,385 (37.9) | 36,957 (37.1) | -0.02 | 25,175 (37.5) | 25,207 (37.5) | 0.00 |
| *Acetaminophen* | 2,940 (0.7) | 1,339 (1.3) | 0.07 | 729 (1.1) | 723 (1.1) | 0.00 |
| *Muscle relaxer* | 86,850 (20.4) | 22,508 (22.6) | 0.05 | 14,719 (21.9) | 14,774 (22.0) | 0.00 |
| *Benzodiazepine* | 96,467 (22.6) | 31,326 (31.4) | 0.20 | 19,178 (28.6) | 18,776 (28.0) | -0.01 |
| *Pregabalin* | 13,064 (3.1) | 8,891 (8.9) | 0.25 | 3,698 (5.5) | 4,190 (6.2) | 0.03 |
| *Anti-psychotics* | 14,314 (3.4) | 5,347 (5.4) | 0.10 | 3,020 (4.5) | 3,074 (4.6) | 0.00 |
| *Selective serotonin/ norepinephrine reuptake inhibitors* | 106,151 (24.9) | 33,308 (33.4) | 0.19 | 20,849 (31.1) | 20,164 (30.0) | -0.02 |
| *Bupropion* | 17,203 (4.0) | 5,914 (5.9) | 0.09 | 3,566 (5.3) | 3,434 (5.1) | -0.01 |
| *Trazodone/nefazodone* | 33,626 (7.9) | 12,301 (12.3) | 0.15 | 7,240 (10.8) | 7,035 (10.5) | -0.01 |
| *Monoamine oxidase inhibitor* | 13,096 (3.1) | 4,979 (5.0) | 0.10 | 2,823 (4.2) | 2,744 (4.1) | -0.01 |
| *Duration of opioid use (mean ± standard deviation)* | 109 ± 158 | 191 ± 193 | 0.46 | 163 ± 197 | 169 ± 189 | 0.03 |
| *Average daily milligram morphine equivalents (MME) from index to qualifying dates (median, IQR)* | 18.8 (6.8, 37.5) | 19.6 (6.8, 40) | 0.11 | 18.8 (6.3, 40) | 18.8 (6.4, 40) | 0.01 |
| Days from index to qualifying (median, IQR) | 4 (0, 12) | 7 (1, 14) | 0.16 | 5 (0, 13) | 5 (1, 13) | 0.04 |
| 0-7 | 161,385 (37.9) | 36,957 (37.1) | 0.16 | 39,793 (59.3) | 39,793 (59.3) | 0.00 |
| 8-30 | 2,940 (0.7) | 1,339 (1.3) |  | 27,340 (40.7) | 27,340 (40.7) |  |
| Complications from index to qualifying dates |  |  |  |  |  |  |
| High acute risk | 596 (0.1) | 136 (0.1) | 0.00 | 13 (0.0) | 13 (0.0) | 0.00 |
| Cardiac arrest | 89 (0.0) | 15 (0.0) | 0.00 | <11 | <11 | 0.00 |
| Intubation | 53 (0.0) | <11 | 0.00 | <11 | <11 | 0.00 |
| Mechanical ventilation | 74 (0.0) | 11 (0.0) | -0.01 | <11 | <11 | 0.00 |
| Cardiopulmonary resuscitation | <11 | <11 | 0.00 | <11 | <11 | 0.00 |
| Respiratory failure | 494 (0.1) | 120 (0.1) | 0.00 | 11 (0.0) | 13 (0.0) | 0.00 |
| Moderate acute risk | 27,192 (6.4) | 6,282 (6.3) | 0.00 | 2,379 (3.5) | 2,379 (3.5) | 0.00 |
| Sepsis | 1,229 (0.3) | 256 (0.3) | -0.01 | 86 (0.1) | 79 (0.1) | 0.00 |
| Pneumonia | 2,199 (0.5) | 530 (0.5) | 0.00 | 186 (0.3) | 169 (0.3) | 0.00 |
| Fluid and electrolyte disorders | 7,287 (1.7) | 1,829 (1.8) | 0.01 | 618 (0.9) | 618 (0.9) | 0.00 |
| Weight loss | 2,443 (0.6) | 855 (0.9) | 0.03 | 207 (0.3) | 270 (0.4) | 0.02 |
| Cancer | 17,578 (4.1) | 3,580 (3.6) | -0.03 | 1,569 (2.3) | 1,557 (2.3) | 0.00 |
| Acute myocardial infarction | 805 (0.2) | 181 (0.2) | 0.00 | 65 (0.1) | 56 (0.1) | 0.00 |
| Paralysis | 803 (0.2) | 159 (0.2) | -0.01 | 83 (0.1) | 52 (0.1) | -0.01 |
| Low acute risk | 23,653 (5.5) | 5,855 (5.9) | 0.01 | 2,045 (3.0) | 2,045 (3.0) | 0.00 |
| Coagulopathy | 2,249 (0.5) | 554 (0.6) | 0.00 | 201 (0.3) | 206 (0.3) | 0.00 |
| Gastrointestinal bleed | 726 (0.2) | 190 (0.2) | 0.00 | 63 (0.1) | 63 (0.1) | 0.00 |
| Cardiac arrhythmias | 20,371 (4.8) | 4,915 (4.9) | 0.01 | 1,741 (2.6) | 1,715 (2.6) | 0.00 |
| Stroke | 2,112 (0.5) | 643 (0.6) | 0.02 | 199 (0.3) | 222 (0.3) | 0.01 |

^a^Index=Date person had first fill of gabapentin, TCA, or duloxetine

IQR: Interquartile range

eFigure 3. Primary analysis Kaplan Meier curves for any-time mortality.

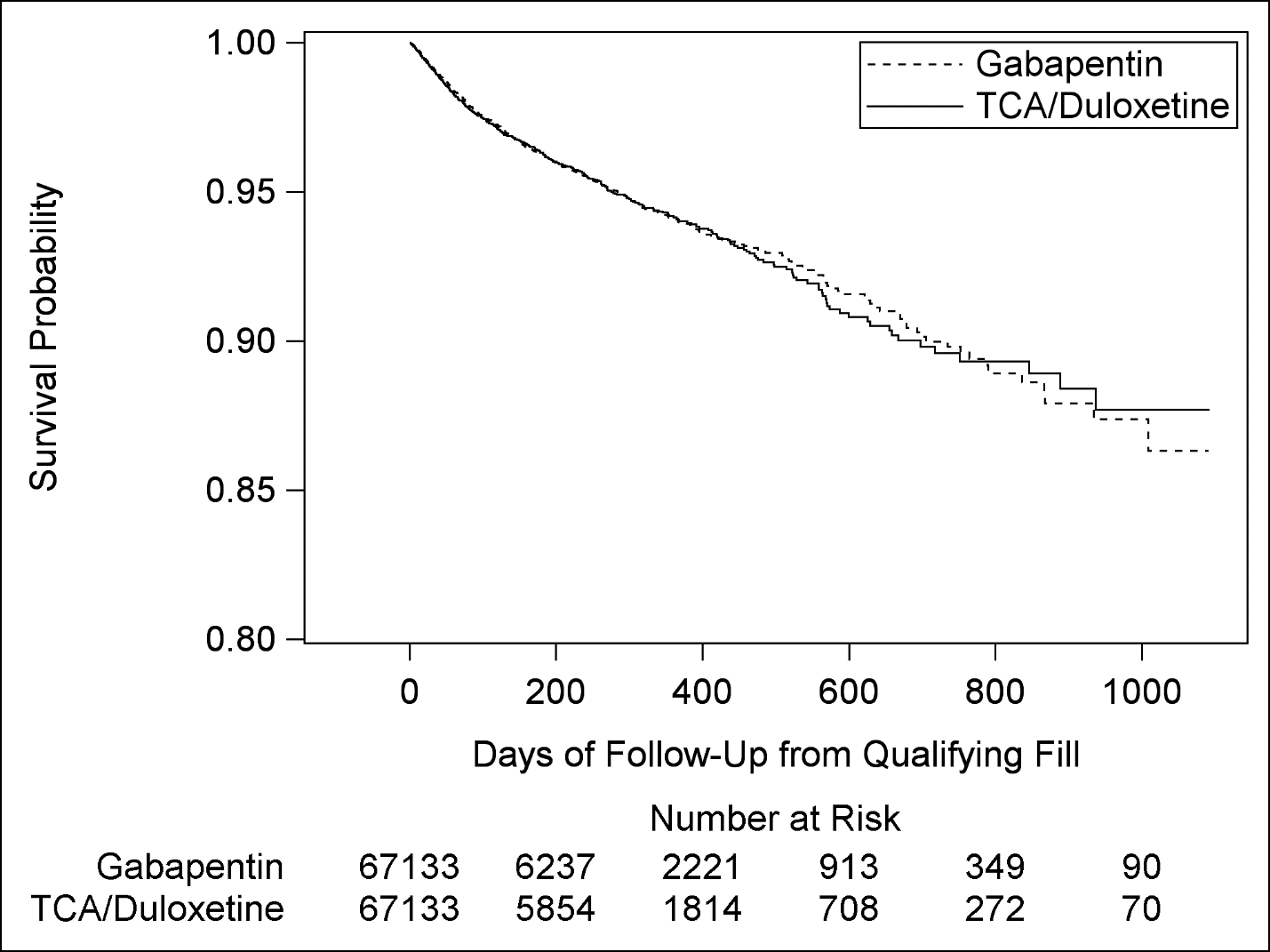

eFigure 4. Primary analysis Kaplan Meier curves for any-time medical complications.

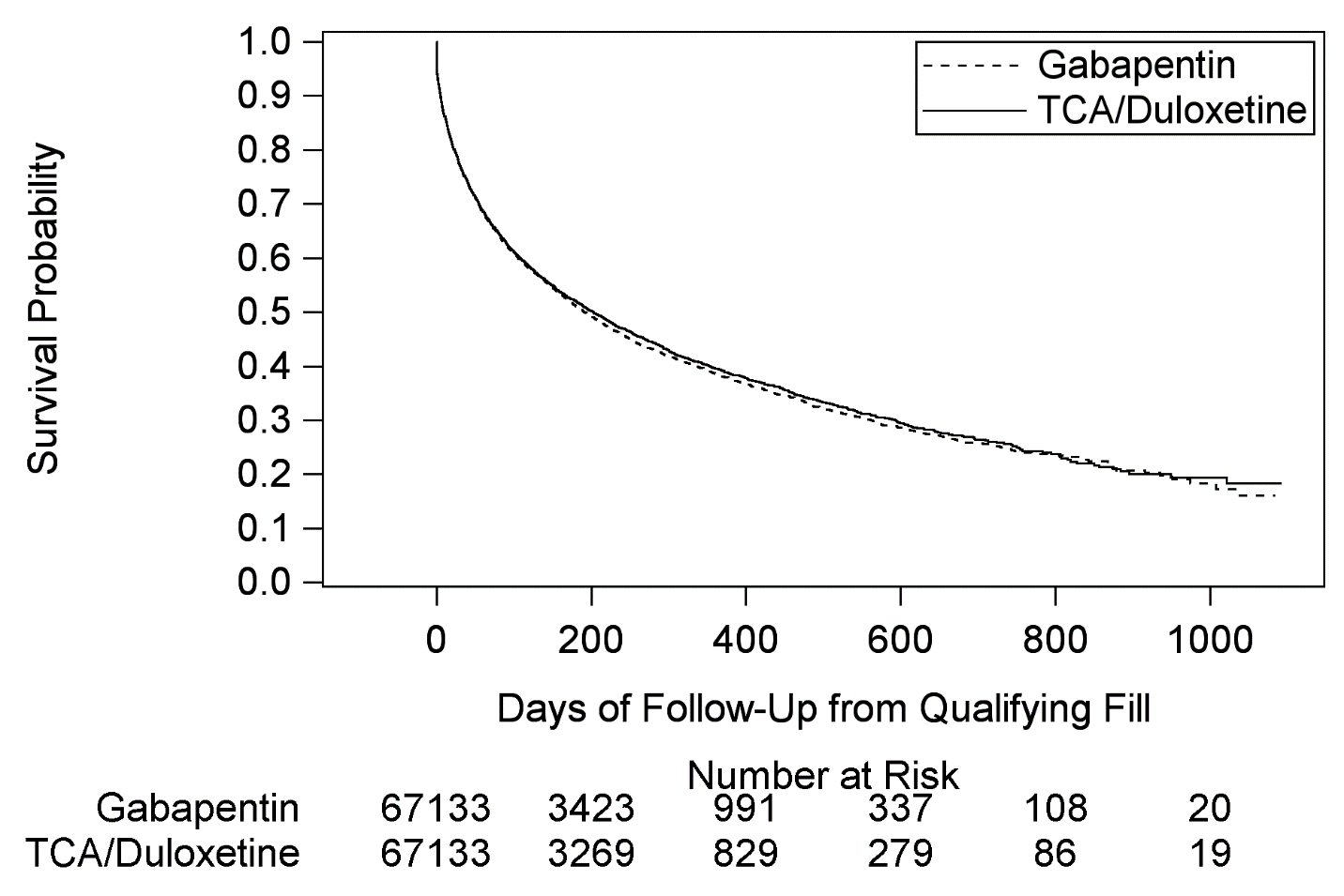

eFigure 5. Primary analysis Cox proportional hazard curves for any-time medical complications.

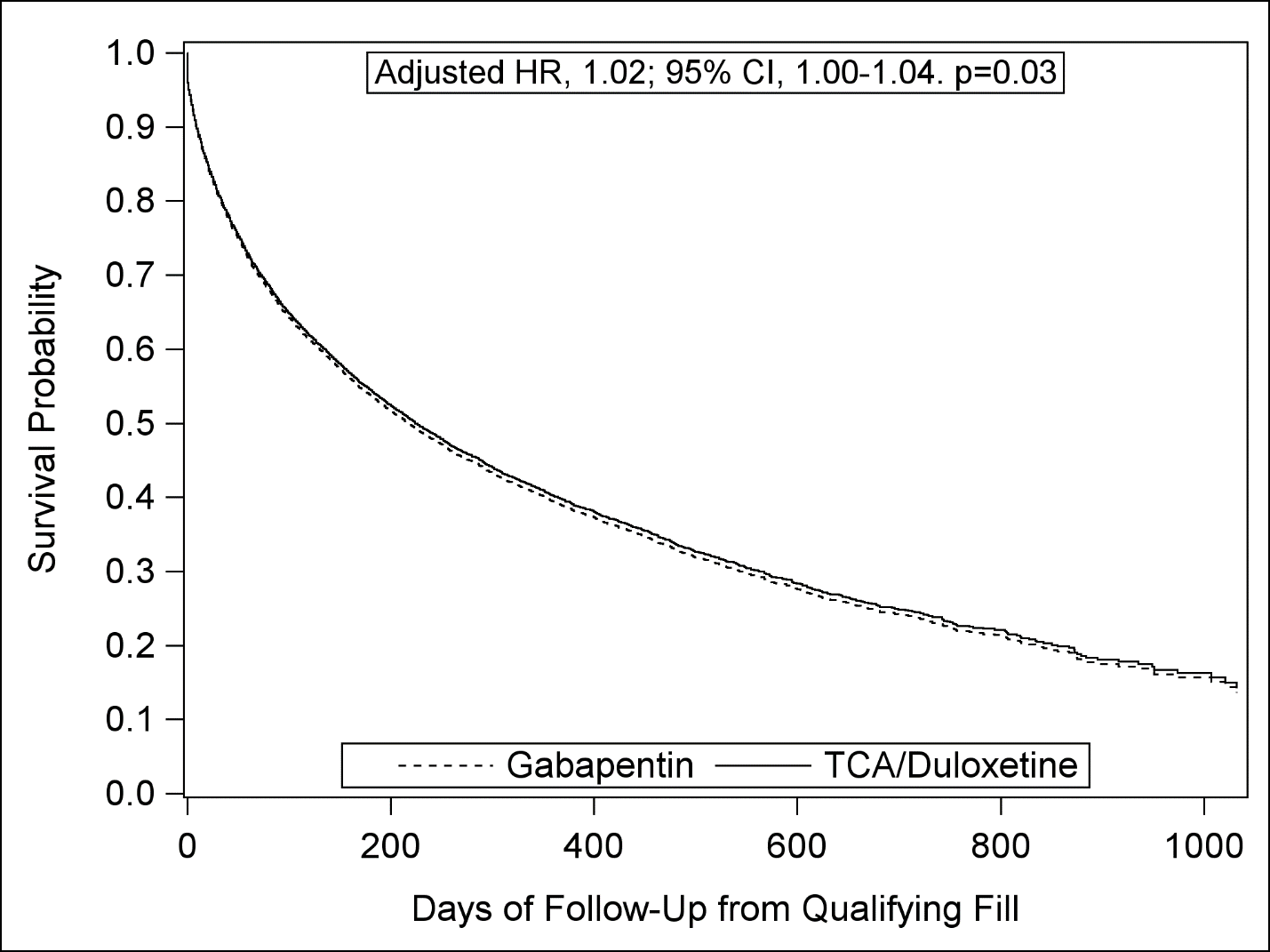

eTable 6. Primary analysis outcomes at 30 days.

| **n (%) unless otherwise indicated** | **Gabapentin n=67,133 (50%)** | **TCA/Duloxetine n=67,133 (50%)** |
| --- | --- | --- |
| **Primary outcome (mortality)** | | |
| Death after qualifying | 453 (0.7) | 466 (0.7) |
| Days to death (median, IQR) | 16 (9, 23) | 15 (8, 22) |
| **Secondary outcome (medical complications)** | | |
| Any complication | 13,931 (20.8) | 13,551 (20.2) |
| Days to first complication (median, IQR) | 6 (0, 15) | 6 (0, 14) |
| Cardiac arrest | 51 (0.1) | 56 (0.1) |
| Intubation | 53 (0.1) | 56 (0.1) |
| Mechanical ventilation | 68 (0.1) | 65 (0.1) |
| Cardiopulmonary resuscitation | 11 (0.0) | 15 (0.0) |
| Respiratory failure | 150 (0.2) | 96 (0.1) |
| Sepsis | 404 (0.6) | 353 (0.5) |
| Pneumonia | 792 (1.2) | 645 (1.0) |
| Fluid and electrolyte disorders | 2,243 (3.3) | 2,246 (3.3) |
| Weight loss | 798 (1.2) | 1,040 (1.5) |
| Cancer | 4,245 (6.3) | 4,029 (6.0) |
| Acute myocardial infarction | 228 (0.3) | 192 (0.3) |
| Paralysis | 204 (0.3) | 188 (0.3) |
| Coagulopathy | 554 (0.8) | 530 (0.8) |
| Gastrointestinal bleed | 177 (0.3) | 199 (0.3) |
| Cardiac arrhythmias | 6,197 (9.2) | 6,000 (8.9) |
| Stroke | 547 (0.8) | 641 (1.0) |

eFigure 6: Primary analysis Kaplan Meier curves for 30-day mortality.

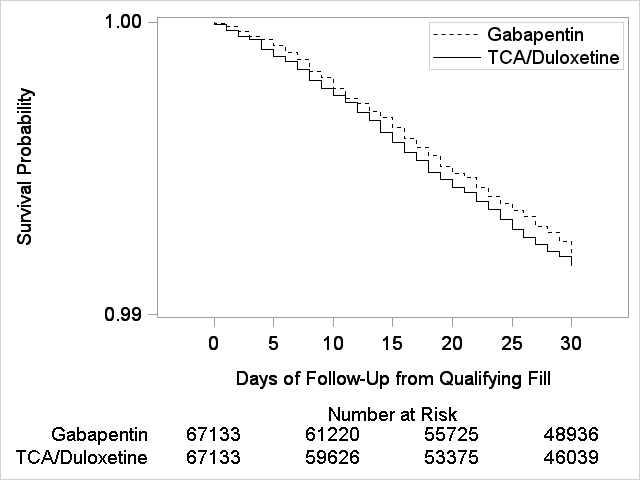

eFigure 7. Primary analysis Cox proportional hazard curves 30-day mortality.

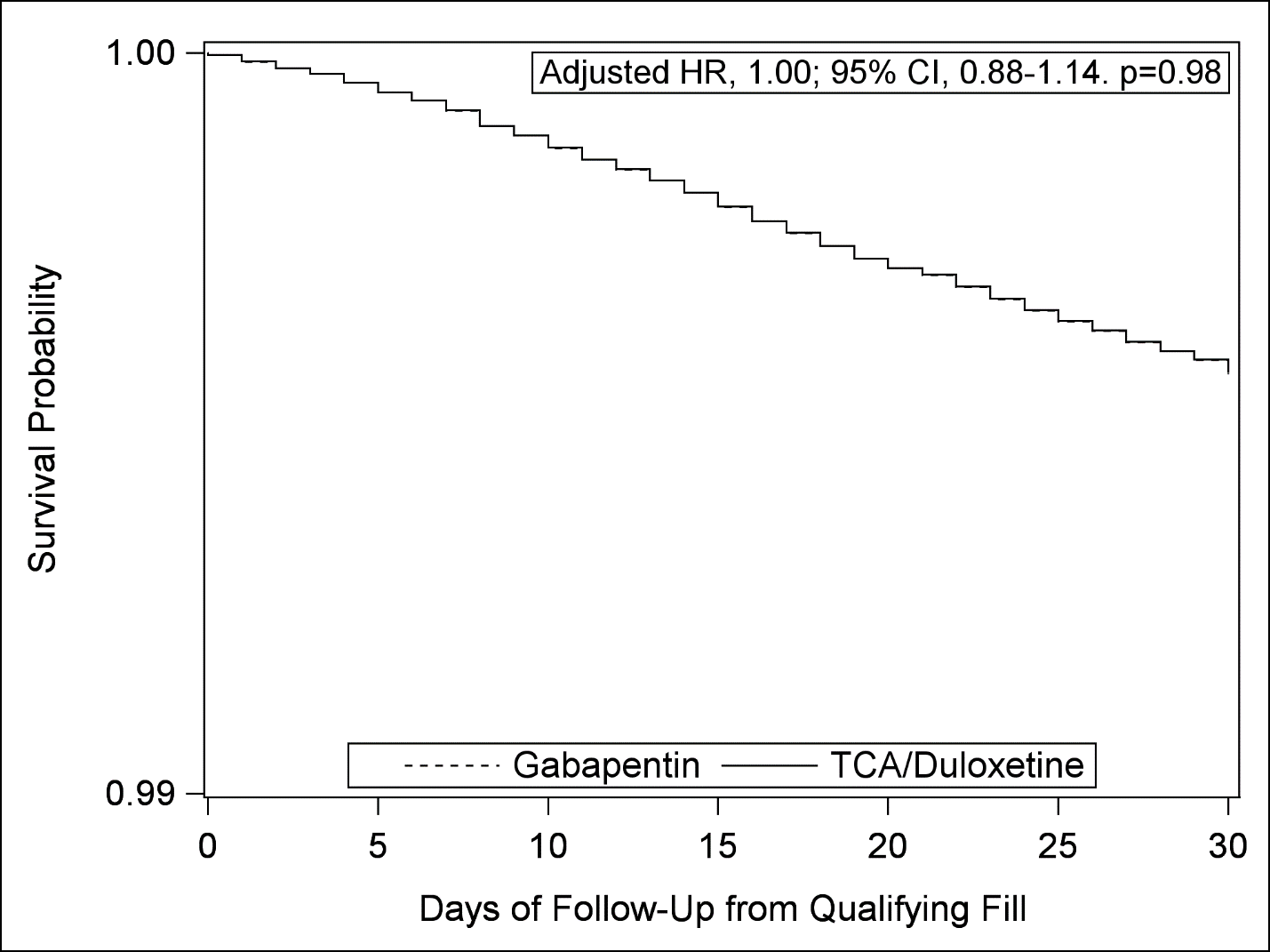

eFigure 8. Primary analysis Kaplan Meier curves for 30-day medical complications.

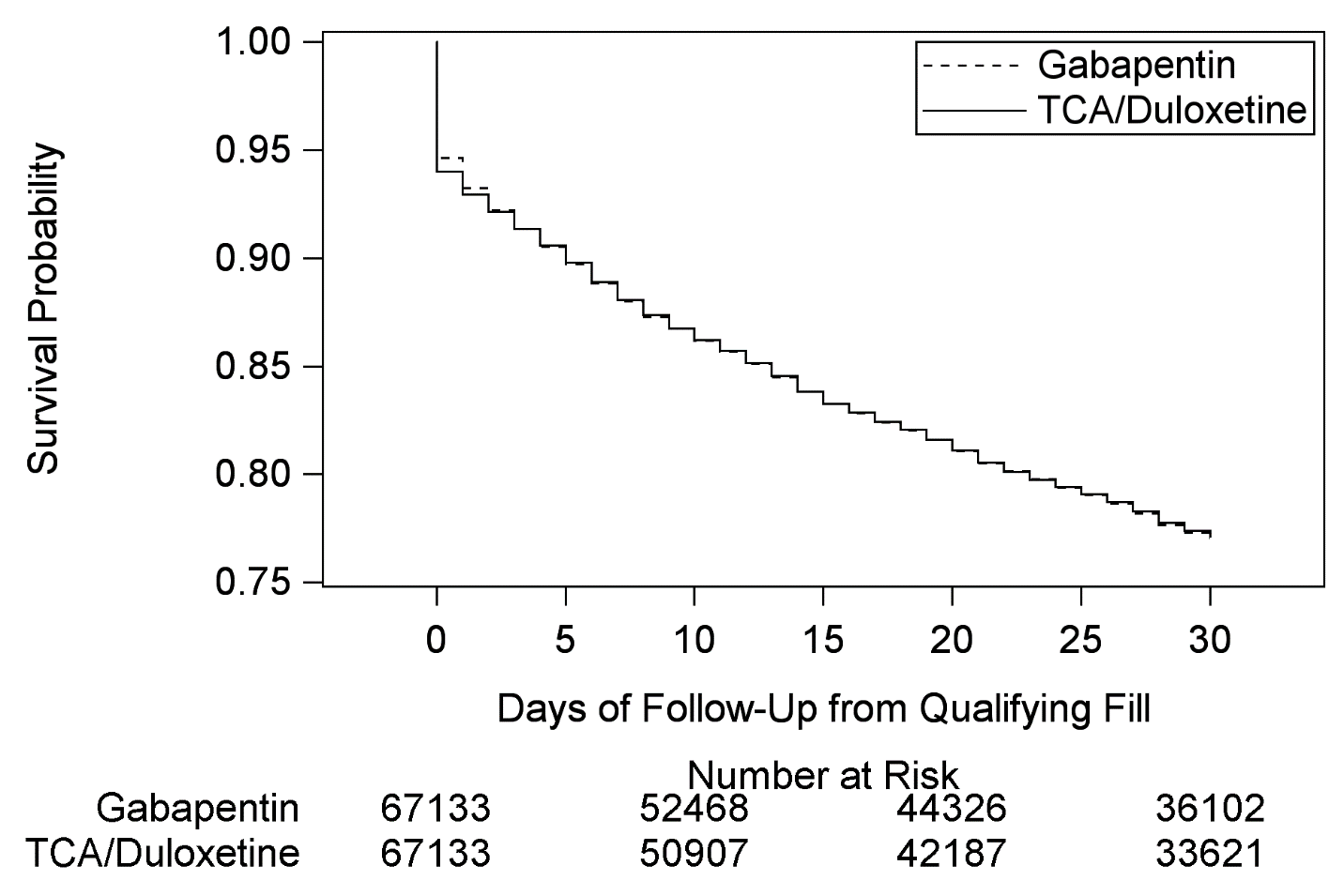

eFigure 9. Primary analysis Cox proportional hazard curves 30-day medical complications.

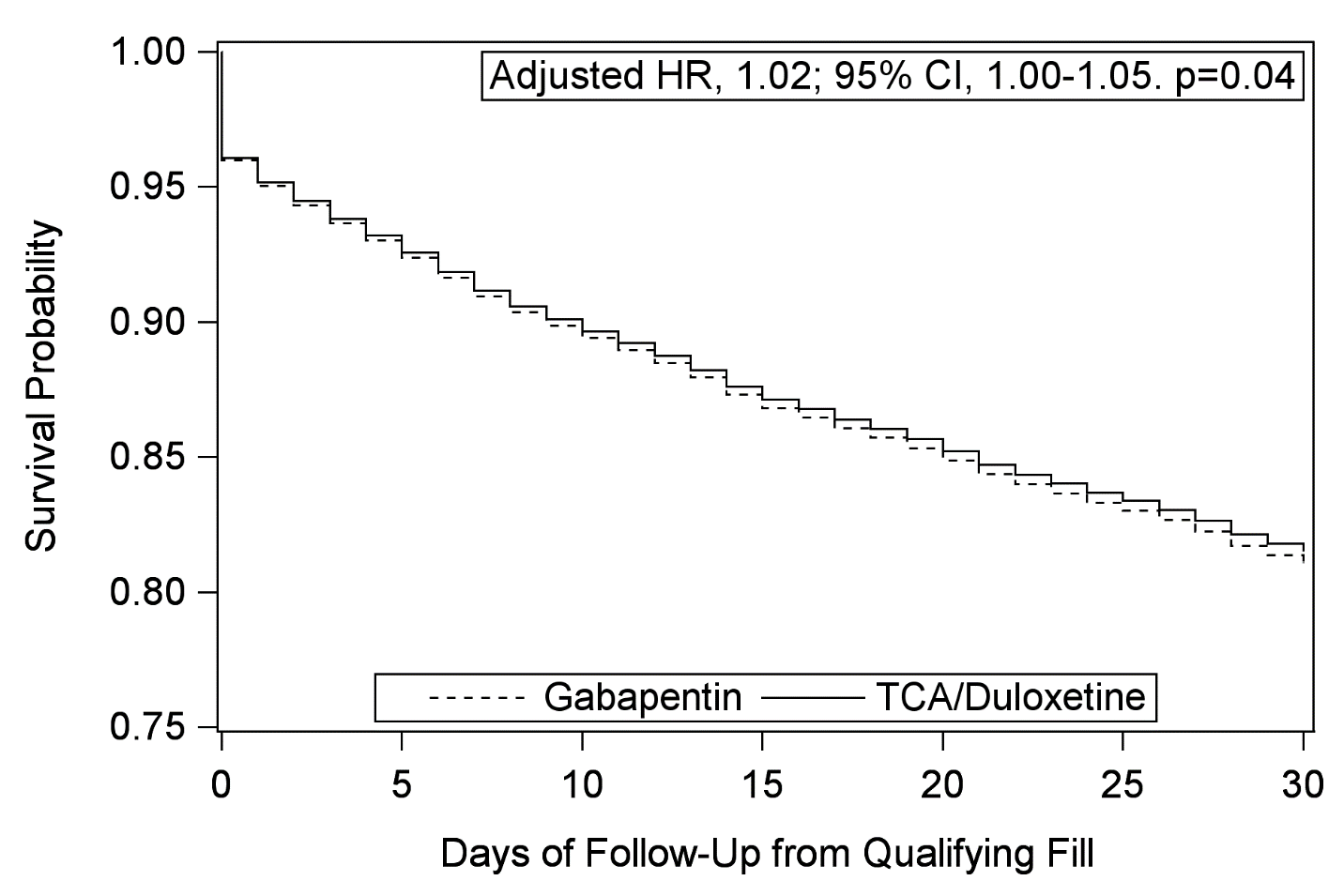

eTable 7. Post hoc analysis comparing hazard ratios for any-time mortality by category of MME on index date replicating aspects of the methods used by Corriere et al.

| Cohort | MME on index date | N | Adjusted^a^ hazard ratio (95% CI) |
| --- | --- | --- | --- |
| - Duloxetine only in active control group.   - Otherwise, same as primary analysis. | None | 34,077 (33.6%) | 0.86 (0.73, 1.01) |
|  | >0 to <50 | 52,993 (52.3%) | 0.98 (0.87, 1.12) |
|  | ≥50 | 14,262 (14.1%) | 0.84 (0.69, 1.03) |
| - Duloxetine only in active control group. - No exact matching on time from index to qualifying dates or medical complications between index and qualifying dates.   - Otherwise, same as primary analysis. | None | 42,575 (33.1%) | 0.81 (0.70, 0.94) |
|  | >0 to <50 | 66,307 (51.6%) | 0.90 (0.80, 1.01) |
|  | ≥50 | 19,700 (15.3%) | 0.90 (0.76, 1.06) |
| - Duloxetine only in active control group. - No exact matching on time from index to qualifying dates or medical complications between index and qualifying dates. - Corriere exclusion criteria (eFigure 2).   - Otherwise, same as primary analysis. | None | 16,083 (31.4) | 0.83 (0.52, 1.34) |
|  | >0 to <50 | 27,789 (54.2) | 1.00 (0.74, 1.34) |
|  | ≥50 | 7409 (14.4) | 0.91 (0.57, 1.45) |

^a^Adjusted for age, sex, race, geographic region, year of index fill, Charlson comorbidity, pain medications in the year prior to index, and number of inpatient and outpatient encounters in the year prior to index. The referent group is the active control (TCA/duloxetine or duloxetine alone).

MME: Milligram morphine equivalents.

eFigure 10. Flow of patients for the secondary analysis.

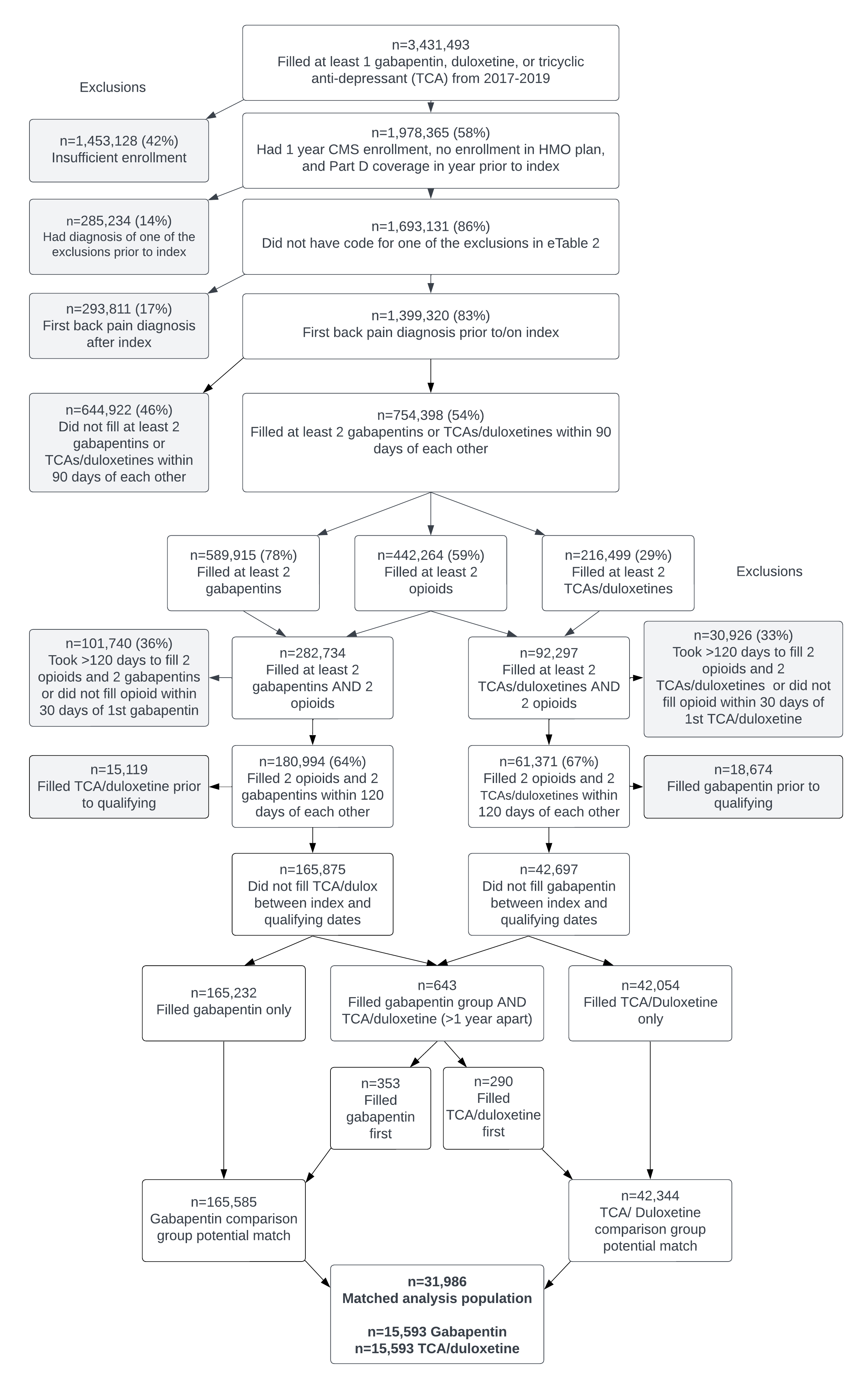

eTable 8. Standardized differences comparing patients who received gabapentin to those who received TCAs/duloxetine in the unmatched and matched populations for the secondary analysis.

|  | **Secondary Analysis Unmatched Population** | | | **Secondary Analysis Matched Population** | | |
| --- | --- | --- | --- | --- | --- | --- |
|  | **Gabapentin n=165,585 (81%) n (%)** | **TCA/Duloxetine n=42,344 (19%)**  **n (%)** | **Standardized difference** | **Gabapentin n=15,993 (50%) n (%)** | **TCA/Duloxetine n=15,993 (50%)**  **n (%)** | **Standardized difference** |
| ***DEMOGRAPHIC VARIABLES*** |  |  |  |  |  |  |
| *Age (median, IQR)* | 73.5 (69.3, 79.4) | 73.0 (68.8, 79.2) | -0.03 | 72.7 (68.9, 78.4) | 72.7 (68.6, 78.9) | 0.02 |
| *Female* | 100,802 (60.9) | 29,237 (69.0) | -0.17 | 10,697 (66.9) | 10,623 (66.4) | 0.01 |
| *Patient race* |  |  |  |  |  |  |
| *Unknown* | 2,067 (1.2) | 381 (0.9) | -0.03 | 169 (1.1) | 170 (1.1) | 0.00 |
| *White* | 142,717 (86.2) | 37,476 (88.5) | 0.07 | 13,938 (87.2) | 13,891 (86.9) | -0.01 |
| *Black* | 14,020 (8.5) | 3,054 (7.2) | -0.05 | 1,318 (8.2) | 1,329 (8.3) | 0.00 |
| *Other* | 1,503 (0.9) | 312 (0.7) | -0.02 | 127 (0.8) | 124 (0.8) | 0.00 |
| *Asian* | 1,746 (1.1) | 275 (0.6) | -0.04 | 115 (0.7) | 122 (0.8) | 0.01 |
| *Hispanic* | 2,588 (1.6) | 534 (1.3) | -0.03 | 220 (1.4) | 236 (1.5) | 0.01 |
| *North American Native* | 944 (0.6) | 312 (0.7) | 0.02 | 106 (0.7) | 121 (0.8) | 0.01 |
| *Region* |  |  |  |  |  |  |
| *New England* | 7,559 (4.6) | 1,640 (3.9) | -0.03 | 589 (3.7) | 640 (4.0) | 0.02 |
| *NY/NJ/PR* | 10,793 (6.5) | 2,313 (5.5) | -0.04 | 903 (5.6) | 953 (6.0) | 0.01 |
| *PA/DE/MD/DC/WV/VA* | 15,202 (9.2) | 3,777 (8.9) | -0.01 | 1,460 (9.1) | 1,481 (9.3) | 0.00 |
| *KY/TN/NC/SC/GA/AL/MS/FL* | 39,953 (24.1) | 10,967 (25.9) | 0.04 | 4,200 (26.3) | 4,018 (25.1) | -0.03 |
| *MN/WI/MI/OH/IN/IL* | 27,747 (16.8) | 7,341 (17.3) | 0.02 | 2,639 (16.5) | 2,694 (16.8) | 0.01 |
| *NM/TX/OK/AR/LA* | 22,762 (13.7) | 5,703 (13.5) | -0.01 | 2,174 (13.6) | 2,167 (13.5) | 0.00 |
| *NE/IA/KS/MO* | 10,175 (6.1) | 2,694 (6.4) | 0.01 | 990 (6.2) | 993 (6.2) | 0.00 |
| *MT/ND/SD/WY/UT/CO* | 5,139 (3.1) | 1,439 (3.4) | 0.02 | 540 (3.4) | 537 (3.4) | 0.00 |
| *CA/HI/NV/AZ/Pac Islands* | 19,665 (11.9) | 4,650 (11.0) | -0.03 | 1,842 (11.5) | 1,848 (11.6) | 0.00 |
| *AK/WA/OR/ID* | 6,590 (4.0) | 1,820 (4.3) | 0.02 | 656 (4.1) | 662 (4.1) | 0.00 |
| *Buy-in from state* | 38,249 (23.1) | 11,855 (28.0) | 0.11 | 4,119 (25.8) | 4,145 (25.9) | 0.00 |
| *Index^a^ year* |  |  |  |  |  |  |
| *2017* | 68,335 (41.3) | 18,179 (42.9) | 0.03 | 6,911 (43.2) | 6,817 (42.6) | -0.01 |
| *2018* | 55,470 (33.5) | 13,810 (32.6) | -0.02 | 5,351 (33.5) | 5,302 (33.2) | -0.01 |
| *2019* | 41,780 (25.2) | 10,355 (24.5) | -0.02 | 3,731 (23.3) | 3,874 (24.2) | 0.02 |
| ***CLINICAL VARIABLES*** |  |  |  |  |  |  |
| *Days from first back pain diagnosis to index (median, IQR)* | 477 (233, 772) | 543 (346, 830) | 0.20 | 532 (330, 812) | 509 (314, 791) | -0.03 |
| Days from most recent back pain diagnosis to index (median, IQR) | 9 (0, 115) | 31 (1, 162) | 0.10 | 12 (0, 128) | 28 (1, 147) | 0.02 |
| *Charlson co-morbidity score (median, IQR)* | 2 (1, 4) | 2 (1, 4) | -0.04 | 2 (1, 4) | 2 (1, 4) | 0.03 |
| *0* | 33,947 (20.5) | 8,468 (20.0) | -0.01 | 3,747 (23.4) | 3,621 (22.6) | -0.02 |
| *1-2* | 56,131 (33.9) | 14,645 (34.6) | 0.01 | 5,818 (36.4) | 5,757 (36.0) | -0.01 |
| *3-4* | 35,772 (21.6) | 9,566 (22.6) | 0.01 | 3,323 (20.8) | 3,415 (21.4) | -0.01 |
| *5+* | 39,735 (24.0) | 9,665 (22.8) | -0.03 | 3,105 (19.4) | 3,200 (20.0) | 0.01 |
| *Diagnoses/procedures (yes/no) in year prior to index* |  |  |  |  |  |  |
| *Cancer (excluding skin cancer)* | 30,480 (18.4) | 6,173 (14.6) | -0.10 | 2,062 (12.9) | 2,133 (13.3) | 0.01 |
| *Metastatic cancer* | 7,769 (4.7) | 1,236 (2.9) | -0.09 | 420 (2.6) | 437 (2.7) | 0.01 |
| *Congestive heart failure* | 36,772 (22.2) | 9,730 (23.0) | 0.02 | 3,018 (18.9) | 3,146 (19.7) | 0.02 |
| *Dementia* | 10,254 (6.2) | 3,874 (9.1) | 0.11 | 1,003 (6.3) | 1,069 (6.7) | 0.02 |
| *Weight loss* | 11,083 (6.7) | 3,454 (8.2) | 0.06 | 1,005 (6.3) | 982 (6.1) | -0.01 |
| *Hemiplegia* | 2,977 (1.8) | 773 (1.8) | 0.00 | 236 (1.5) | 236 (1.5) | 0.00 |
| *Alcohol abuse* | 995 (0.6) | 248 (0.6) | 0.00 | 73 (0.5) | 103 (0.6) | 0.03 |
| *Cardiac arrhythmias* | 54,630 (33.0) | 13,785 (32.6) | -0.01 | 4,188 (26.2) | 4,390 (27.4) | 0.03 |
| *Chronic pulmonary disease* | 53,531 (32.3) | 14,900 (35.2) | 0.06 | 4,998 (31.3) | 5,173 (32.3) | 0.02 |
| *Coagulopathy* | 8,963 (5.4) | 2,024 (4.8) | -0.03 | 621 (3.9) | 665 (4.2) | 0.01 |
| *Diabetes with complications* | 38,906 (23.5) | 9,336 (22.0) | -0.03 | 3,428 (21.4) | 3,533 (22.1) | 0.02 |
| *Deficiency anemia* | 41,415 (25.0) | 11,132 (26.3) | 0.03 | 3,722 (23.3) | 3,777 (23.6) | 0.01 |
| *Fluid/electrolyte disorders* | 31,298 (18.9) | 8,558 (20.2) | 0.03 | 2,610 (16.3) | 2,679 (16.8) | 0.01 |
| *Liver disease* | 12,644 (7.6) | 3,229 (7.6) | 0.00 | 1,105 (6.9) | 1,131 (7.1) | 0.01 |
| *Peripheral vascular disease* | 41,131 (24.8) | 10,371 (24.5) | -0.01 | 3,660 (22.9) | 3,653 (22.8) | 0.00 |
| *Pulmonary circulation disorders* | 9,739 (5.9) | 2,615 (6.2) | 0.01 | 792 (5.0) | 800 (5.0) | 0.00 |
| *HIV/AIDS* | 350 (0.2) | 96 (0.2) | 0.00 | 27 (0.2) | 31 (0.2) | 0.01 |
| *Hypertension* | 132,938 (80.3) | 33,757 (79.7) | -0.01 | 12,490 (78.1) | 12,571 (78.6) | 0.01 |
| *Myocardial infarction* | 5,177 (3.1) | 1,289 (3.0) | 0.00 | 350 (2.2) | 435 (2.7) | 0.03 |
| *Rheumatic disease* | 12,122 (7.3) | 4,244 (10.0) | 0.10 | 1,386 (8.7) | 1,382 (8.6) | 0.00 |
| *Peptic ulcer disease* | 4,206 (2.5) | 1,199 (2.8) | 0.02 | 412 (2.6) | 391 (2.4) | -0.01 |
| *Diabetes without complications* | 57,729 (34.9) | 13,870 (32.8) | -0.04 | 5,156 (32.2) | 5,264 (32.9) | 0.01 |
| *Opioid use disorder* | 28,246 (17.1) | 11,545 (27.3) | 0.25 | 3,990 (24.9) | 3,902 (24.4) | -0.01 |
| *Parkinson’s disease* | 2,771 (1.7) | 810 (1.9) | 0.02 | 283 (1.8) | 270 (1.7) | -0.01 |
| *Obesity* | 31,013 (18.7) | 7,863 (18.6) | 0.00 | 2,887 (18.1) | 2,945 (18.4) | 0.01 |
| *Insomnia* | 18,604 (11.2) | 6,883 (16.3) | 0.15 | 2,120 (13.3) | 2,127 (13.3) | 0.00 |
| *Sleep apnea* | 23,111 (14.0) | 6,214 (14.7) | 0.02 | 2,157 (13.5) | 2,166 (13.5) | 0.00 |
| *Renal failure* | 32,643 (19.7) | 8,490 (20.1) | 0.01 | 2,853 (17.8) | 2,939 (18.4) | 0.01 |
| *Electroconvulsive therapy* | 21 (0.0) | 19 (0.0) | 0.02 | <11 | <11 | 0.00 |
| *Therapeutic repetitive transcranial magnetic stimulation treatment* | 25 (0.0) | 20 (0.0) | 0.02 | <11 | <11 | 0.00 |
| *Spine-related neuropathic pain* | 98,286 (59.4) | 21,026 (49.7) | -0.20 | 8,658 (54.1) | 8,869 (55.5) | 0.03 |
| *Number of inpatient admissions (median, IQR)* | 0 (0, 1) | 0 (0, 1) | -0.04 | 0 (0, 1) | 0 (0, 1) | -0.03 |
| *Number of outpatient encounters (median, IQR)* | 24 (15, 38) | 25 (15, 39) | 0.02 | 23 (14, 36) | 24 (14, 37) | 0.02 |
| *Number of unique dates with generalized anxiety disorder diagnoses (median, IQR)* | 0 (0, 0) | 0 (0, 0) | 0.09 | 0 (0, 0) | 0 (0, 0) | 0.03 |
| *Number of unique dates with post-traumatic stress disorder diagnoses (median, IQR)* | 0 (0, 0) | 0 (0, 0) | 0.03 | 0 (0, 0) | 0 (0, 0) | 0.04 |
| *Number of unique dates with back pain diagnoses (median, IQR)* | 4 (1, 9) | 4 (1, 10) | 0.02 | 4 (1, 10) | 4 (1, 10) | 0.03 |
| *Number of unique dates with non-spine neuropathy diagnoses (median, IQR)* | 0 (0, 0) | 0 (0, 0) | 0.05 | 0 (0, 0) | 0 (0, 0) | 0.00 |
| *Number of unique dates with depression diagnoses (median, IQR)* | 0 (0, 0) | 0 (0, 1) | 0.17 | 0 (0, 1) | 0 (0, 1) | 0.02 |
| *Medication fills in year prior to index* |  |  |  |  |  |  |
| *NSAID* | 63,805 (38.5) | 15,783 (37.3) | -0.03 | 6,231 (39.0) | 6,290 (39.3) | 0.01 |
| *Acetaminophen* | 1,267 (0.8) | 580 (1.4) | 0.06 | 191 (1.2) | 174 (1.1) | -0.01 |
| *Muscle relaxer* | 37,665 (22.7) | 10,258 (24.2) | 0.03 | 3,858 (24.1) | 3,905 (24.4) | 0.01 |
| *Benzodiazepine* | 41,770 (25.2) | 14,030 (33.1) | 0.17 | 4,805 (30.0) | 4,673 (29.2) | -0.02 |
| *Pregabalin* | 6,427 (3.9) | 4,568 (10.8) | 0.27 | 1,013 (6.3) | 1,183 (7.4) | 0.04 |
| *Anti-psychotics* | 7,228 (4.4) | 2,884 (6.8) | 0.11 | 853 (5.3) | 899 (5.6) | 0.01 |
| *Selective serotonin/ norepinephrine reuptake inhibitors* | 46,765 (28.2) | 16,453 (38.9) | 0.23 | 5,487 (34.3) | 5,416 (33.9) | -0.01 |
| *Bupropion* | 7,510 (4.5) | 2,877 (6.8) | 0.10 | 895 (5.6) | 935 (5.8) | 0.01 |
| *Trazodone/nefazodone* | 15,949 (9.6) | 6,079 (14.4) | 0.15 | 1,928 (12.1) | 1,921 (12.0) | 0.00 |
| *Monoamine oxidase inhibitor* | 6,356 (3.8) | 2,512 (5.9) | 0.10 | 753 (4.7) | 745 (4.7) | 0.00 |
| *Duration of opioid use (mean ± standard deviation)* | 153 ± 177 | 237 ± 197 | 0.45 | 222 ± 213 | 217 ± 202 | -0.02 |
| *Average daily milligram morphine equivalents (MME) from index to qualifying dates (median, IQR)* | 15.3 (7.9, 30.8) | 20.1 (9.7, 42.6) | 0.21 | 19.0 (8.9, 41.3) | 19.0 (8.9, 41.7) | 0.00 |
| Days from index to qualifying (median, IQR) | 47 (33, 61) | 50 (37, 63) | 0.20 | 51 (38, 63) | 50 (38, 63) | 0.03 |
| 0-30 | 33,627 (20.3) | 4,978 (11.8) | -0.23 | 1,699 (10.6) | 1,699 (10.6) | 0.00 |
| 31-45 | 45,441 (27.4) | 11,938 (28.2) | 0.02 | 4,628 (28.9) | 4,628 (28.9) | 0.00 |
| 46-60 | 42,891 (25.9) | 12,793 (30.2) | 0.10 | 4,954 (31.0) | 4,954 (31.0) | 0.00 |
| 61-120 | 43,626 (26.3) | 12,635 (29.8) | 0.08 | 4,712 (29.5) | 4,712 (29.5) | 0.00 |
| Complications from index to qualifying dates |  |  |  |  |  |  |
| High acute risk | 1,332 (0.8) | 235 (0.6) | -0.03 | 15 (0.1) | 15 (0.1) | 0.00 |
| Cardiac arrest | 179 (0.1) | 27 (0.1) | -0.02 | <11 | 0 | 0.00 |
| Intubation | 161 (0.1) | 19 (0.0) | -0.02 | <11 | <11 | 0.00 |
| Mechanical ventilation | 214 (0.1) | 27 (0.1) | -0.02 | <11 | <11 | 0.01 |
| Cardiopulmonary resuscitation | 14 (0.0) | <11 | 0.00 | <11 | <11 | 0.01 |
| Respiratory failure | 1,133 (0.7) | 205 (0.5) | -0.03 | 14 (0.1) | 12 (0.1) | 0.00 |
| Moderate acute risk | 33,516 (20.2) | 7,509 (17.7) | -0.06 | 1,615 (10.1) | 1,615 (10.1) | 0.00 |
| Sepsis | 2,567 (1.6) | 517 (1.2) | -0.03 | 110 (0.7) | 90 (0.6) | -0.02 |
| Pneumonia | 4,297 (2.6) | 960 (2.3) | -0.02 | 195 (1.2) | 185 (1.2) | -0.01 |
| Fluid and electrolyte disorders | 12,313 (7.4) | 2,900 (6.8) | -0.02 | 547 (3.4) | 564 (3.5) | 0.01 |
| Weight loss | 4,266 (2.6) | 1,394 (3.3) | 0.04 | 210 (1.3) | 267 (1.7) | 0.03 |
| Cancer | 18,671 (11.3) | 3,469 (8.2) | -0.10 | 936 (5.9) | 919 (5.7) | 0.00 |
| Acute myocardial infarction | 1,401 (0.8) | 283 (0.7) | -0.02 | 60 (0.4) | 52 (0.3) | -0.01 |
| Paralysis | 1,366 (0.8) | 246 (0.6) | -0.03 | 64 (0.4) | 40 (0.3) | -0.03 |
| Low acute risk | 31,730 (19.2) | 7,532 (17.8) | -0.04 | 1,426 (8.9) | 1,426 (8.9) | 0.00 |
| Coagulopathy | 3,668 (2.2) | 746 (1.8) | -0.03 | 159 (1.0) | 154 (1.0) | 0.00 |
| Gastrointestinal bleed | 1,288 (0.8) | 316 (0.7) | 0.00 | 55 (0.3) | 61 (0.4) | 0.01 |
| Cardiac arrhythmias | 27,157 (16.4) | 6,416 (15.2) | -0.03 | 1,217 (7.6) | 1,220 (7.6) | 0.00 |
| Stroke | 3,211 (1.9) | 896 (2.1) | 0.01 | 142 (0.9) | 170 (1.1) | 0.02 |

^a^Index=Date person had first fill of gabapentin, TCA, or duloxetine

IQR: Interquartile range

eTable 9. Secondary analysis outcomes at any time and reasons for end of follow-up.

| **n (%) unless otherwise indicated** | **Gabapentin n=15,993 (50%)** | **TCA/Duloxetine n=15,993 (50%)** |
| --- | --- | --- |
| **Primary outcome (mortality)** | | |
| Death after qualifying | 283 (1.8) | 262 (1.6) |
| Days to death (median, IQR) | 83 (34, 261) | 57 (27, 153) |
| **Secondary outcome (medical complications)** | | |
| Any complication | 4,644 (29.0) | 4,401 (27.5) |
| Days to first complication (median, IQR) | 26 (8, 72) | 25 (7, 70) |
| Cardiac arrest | 30 (0.2) | 27 (0.2) |
| Intubation | 27 (0.2) | 22 (0.1) |
| Mechanical ventilation | 34 (0.2) | 26 (0.2) |
| Cardiopulmonary resuscitation | <11 | <11 |
| Respiratory failure | 61 (0.4) | 50 (0.3) |
| Sepsis | 210 (1.3) | 172 (1.1) |
| Pneumonia | 380 (2.4) | 321 (2.0) |
| Fluid and electrolyte disorders | 973 (6.1) | 933 (5.8) |
| Weight loss | 320 (2.0) | 331 (2.1) |
| Cancer | 1,085 (6.8) | 1,019 (6.4) |
| Acute myocardial infarction | 113 (0.7) | 98 (0.6) |
| Paralysis | 75 (0.5) | 50 (0.3) |
| Coagulopathy | 182 (1.1) | 183 (1.1) |
| Gastrointestinal bleed | 72 (0.5) | 83 (0.5) |
| Cardiac arrhythmias | 2,010 (12.6) | 1,893 (11.8) |
| Stroke | 214 (1.3) | 194 (1.2) |
| **Reasons for follow-up end** |  |  |
| Disenrolled from Medicare/end of study | 3,225 (20.2) | 3,081 (19.3) |
| No opioid refill >45 days | 7,146 (44.7) | 6,457 (40.4) |
| Pregabalin fill | 459 (2.9) | 979 (6.1) |
| Benzodiazepine fill | 3,222 (20.1) | 3,101 (19.4) |
| TCA/duloxetine fill in gabapentin group | 524 (3.3) | -- |
| Gabapentin fill in TCA/duloxetine group | -- | 910 (5.7) |
| No gabapentin refill >180 days | 1,134 (7.1) | -- |
| No TCA/duloxetine refill >180 days | -- | 1,203 (7.5) |
| Death | 283 (1.8) | 262 (1.6) |

eFigure 11. Secondary analysis Kaplan Meier curves for any-time mortality.

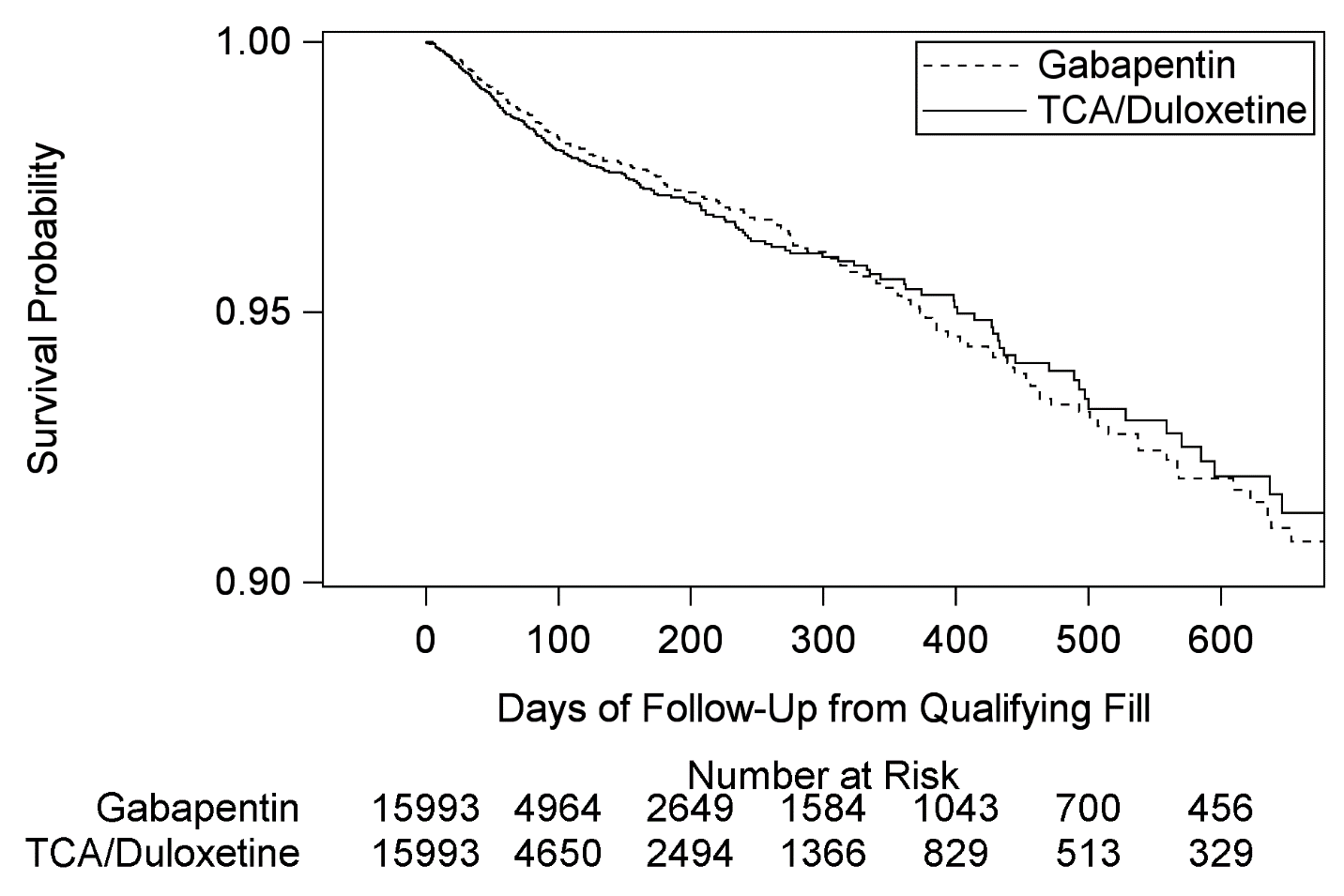

eFigure 12. Secondary analysis Cox proportional hazard curves for any-time mortality.

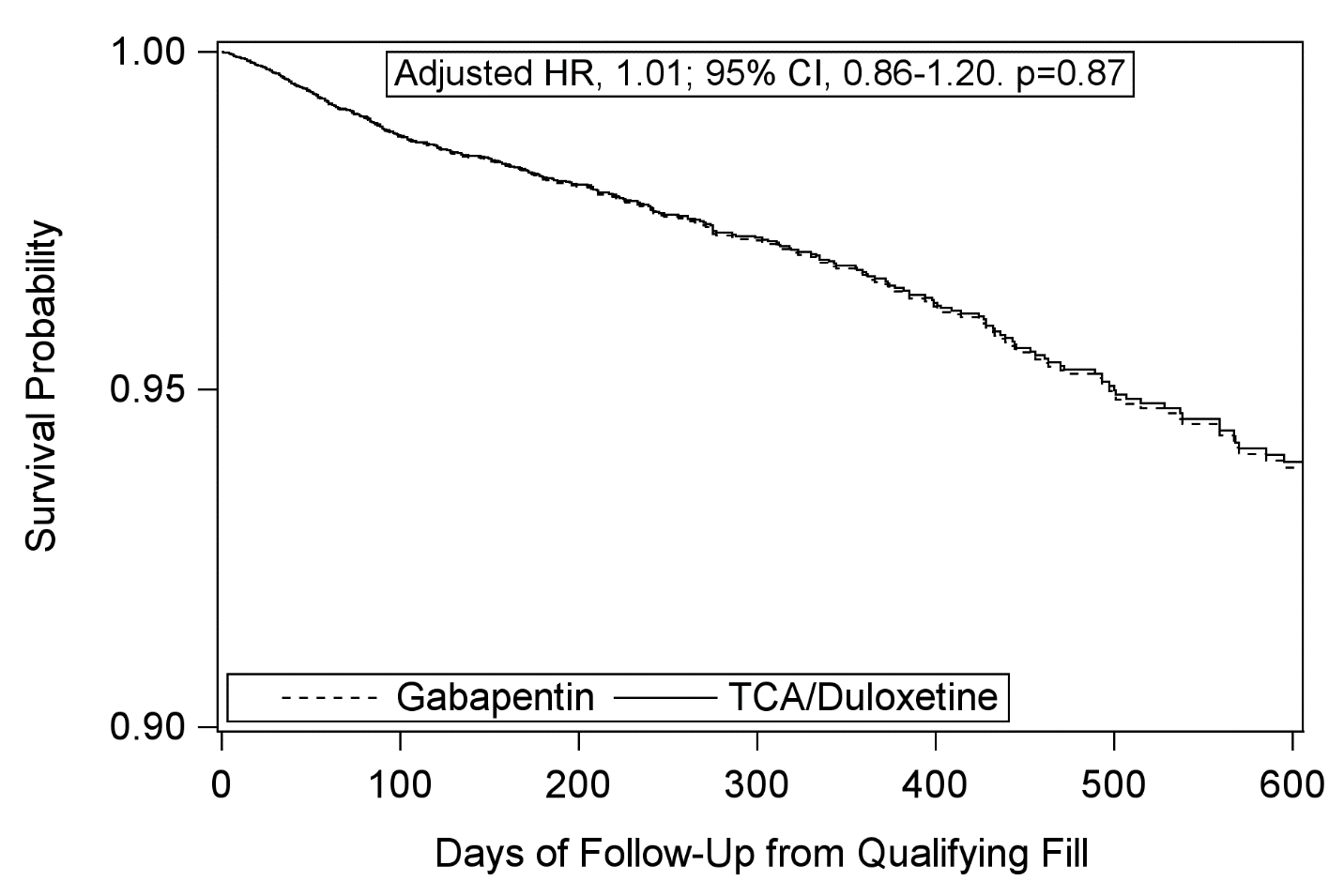

eTable 10. Secondary analysis: *post hoc* analysis comparing hazard ratios for any-time mortality among quartiles of average daily MME from fills from index to qualifying dates.

| MME level | N | Adjusted^a^ hazard ratio (95% CI) |
| --- | --- | --- |
| >0 to 8.16 | 7202 (22.5) | 0.85 (0.56, 1.29) |
| ≥8.17 to 16.0 | 7062 (22.1) | 0.77 (0.52, 1.16) |
| ≥16.0 to 32.6 | 7495 (23.4) | 1.33 (0.94, 1.89) |
| ≥32.6 | 10,227 (32.0) | 0.98 (0.75, 1.28) |

^a^Adjusted for age, sex, race, geographic region, year of index fill, Charlson comorbidity, pain medications in the year prior to index, and number of inpatient and outpatient encounters in the year prior to index.

MME: Milligram morphine equivalents.

eFigure 13. Secondary analysis Kaplan Meier curves for any-time medical complications.

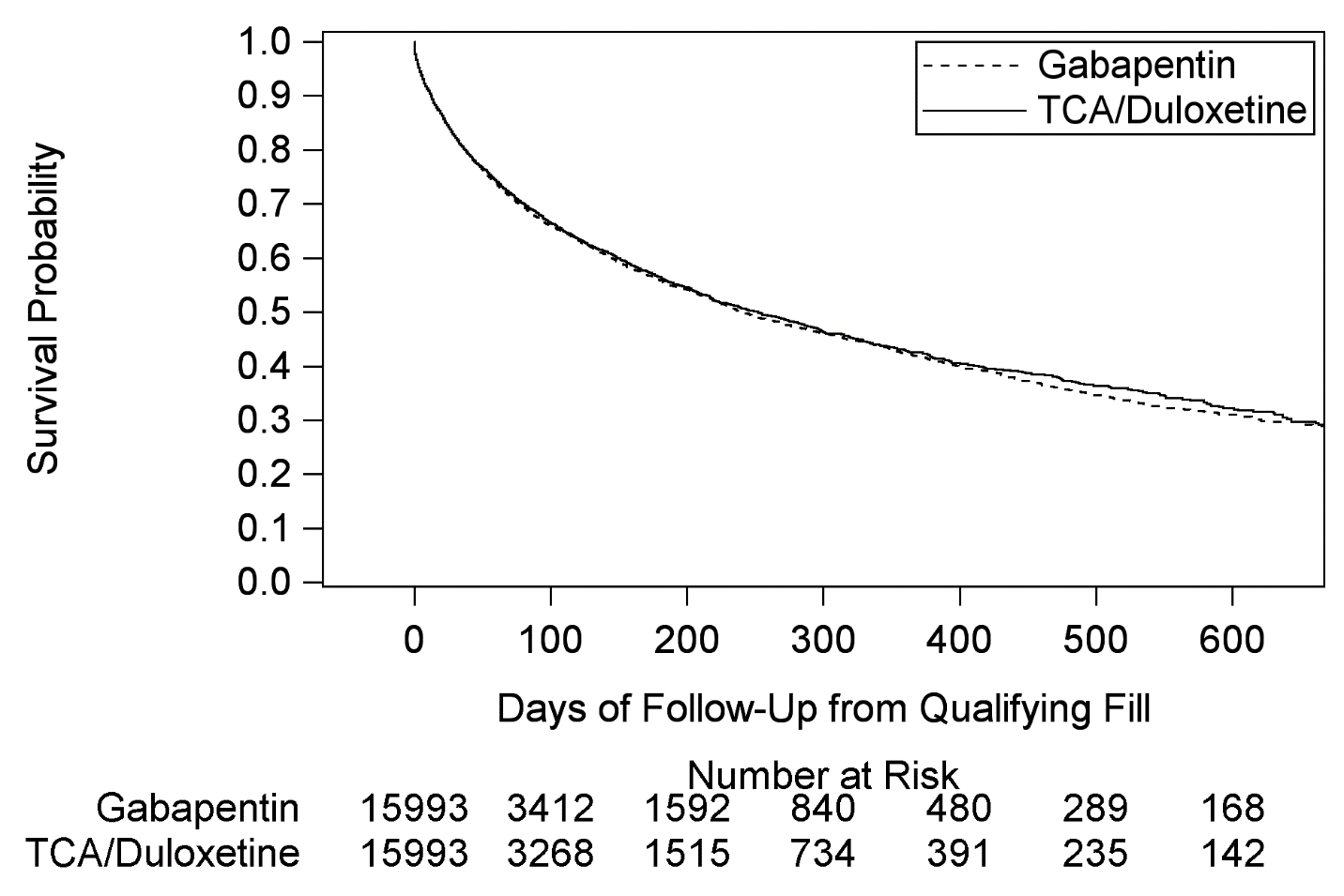

eFigure 14. Secondary analysis Cox proportional hazard curves for any-time medical complications.

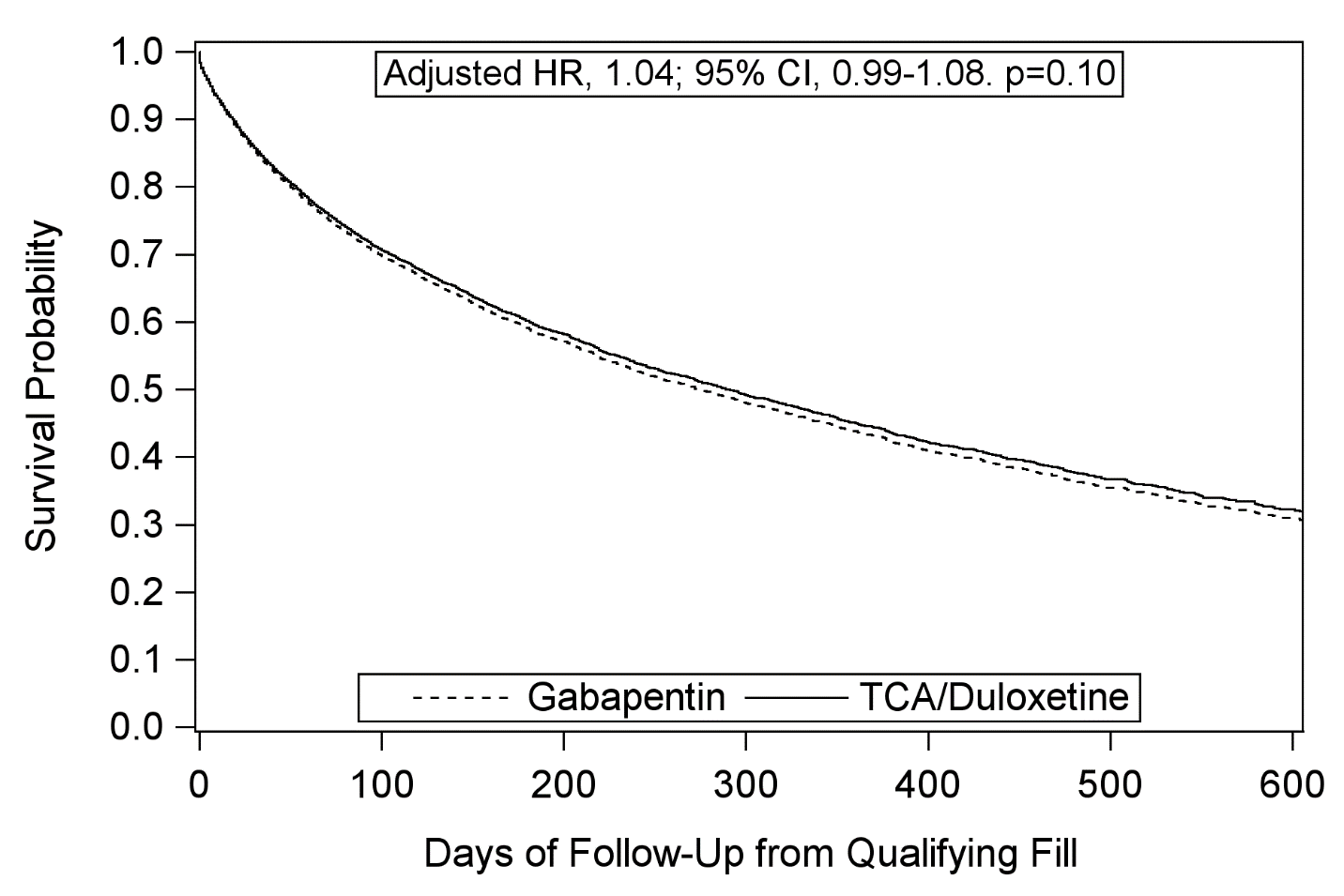

eTable 11. Secondary analysis outcomes at 30-days.

| **n (%) unless otherwise indicated** | **Gabapentin n=15,993 (50%)** | **TCA/Duloxetine n=15,993 (50%)** |
| --- | --- | --- |
| **Primary outcome (mortality)** | | |
| Death after qualifying | 61 (0.4) | 73 (0.5) |
| Days to death (median, IQR) | 16 (7, 24) | 18 (10, 24) |
| **Secondary outcome (medical complications)** | | |
| Any complication | 2,511 (15.7) | 2,456 (15.4) |
| Days to first complication (median, IQR) | 9 (2, 18) | 8 (2, 18) |
| Cardiac arrest | <11 | <11 |
| Intubation | 12 (0.1) | <11 |
| Mechanical ventilation | 14 (0.1) | 107 (1) |
| Cardiopulmonary resuscitation | <11 | 0 |
| Respiratory failure | 26 (0.2) | 21 (0.1) |
| Sepsis | 80 (0.5) | 78 (0.5) |
| Pneumonia | 151 (0.9) | 147 (0.9) |
| Fluid and electrolyte disorders | 434 (2.7) | 427 (2.7) |
| Weight loss | 132 (0.8) | 170 (1.1) |
| Cancer | 781 (4.9) | 717 (4.5) |
| Acute myocardial infarction | 48 (0.3) | 41 (0.3) |
| Paralysis | 36 (0.2) | 27 (0.2) |
| Coagulopathy | 103 (0.6) | 98 (0.6) |
| Gastrointestinal bleed | 32 (0.2) | 44 (0.3) |
| Cardiac arrhythmias | 1,101 (6.9) | 1,075 (6.7) |
| Stroke | 104 (0.7) | 102 (0.6) |

eFigure 15. Secondary analysis Kaplan Meier curves for 30-day mortality.

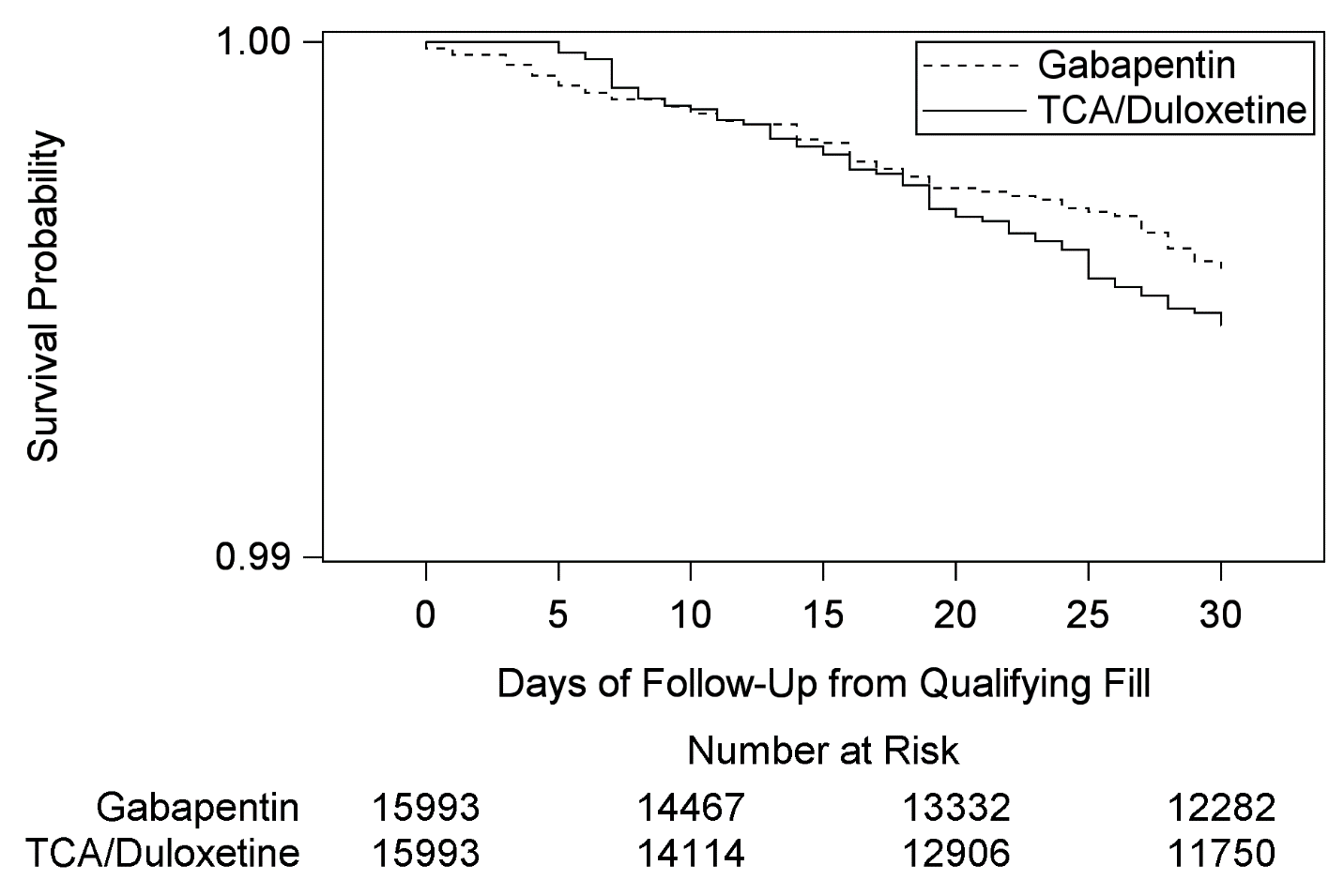

eFigure 16. Secondary analysis Cox proportional hazard curves for 30-day mortality.

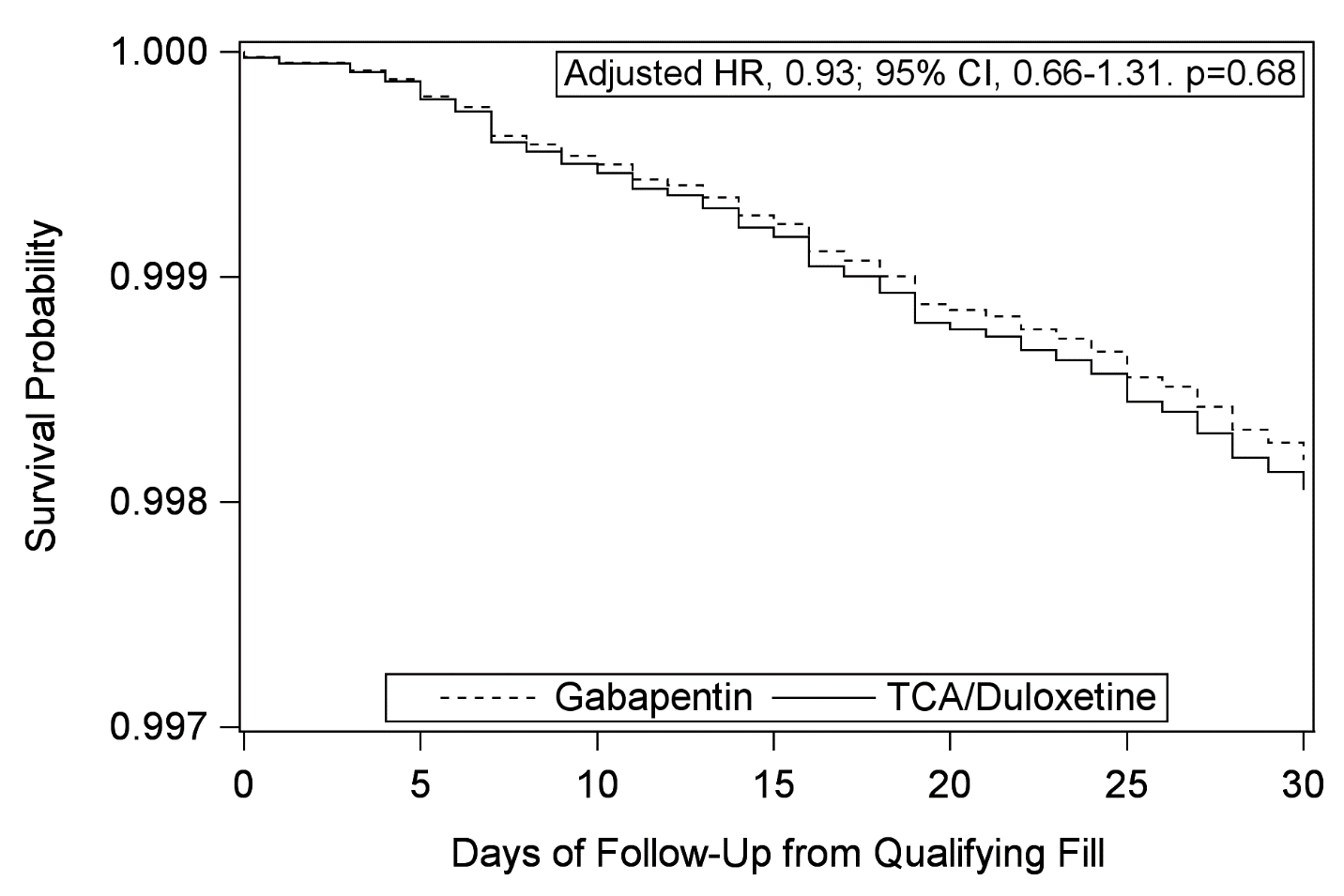

eFigure 17. Secondary analysis Kaplan Meier curves for 30-day medical complications.

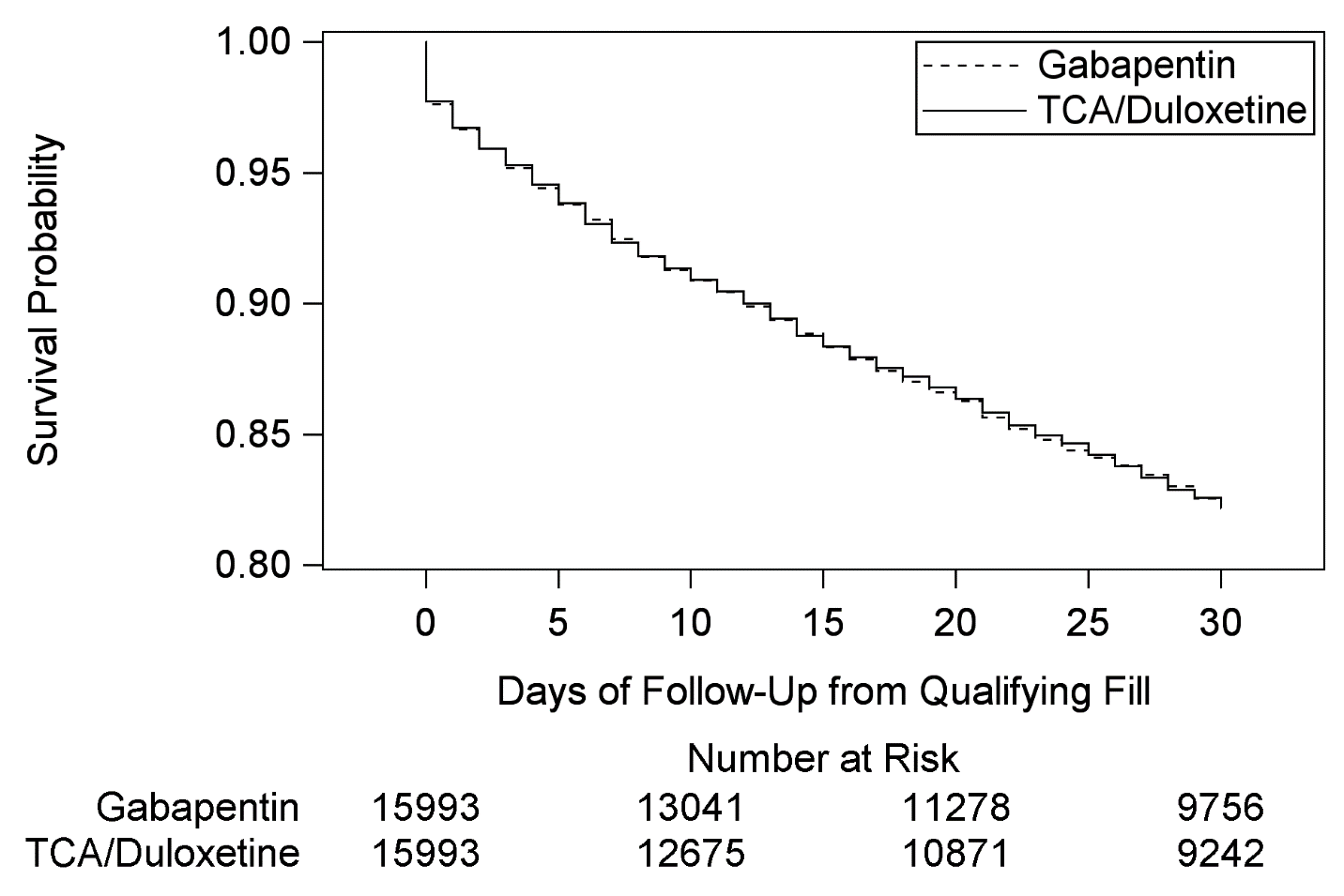

eFigure 18. Secondary analysis Cox proportional hazard curves for 30-day medical complications.

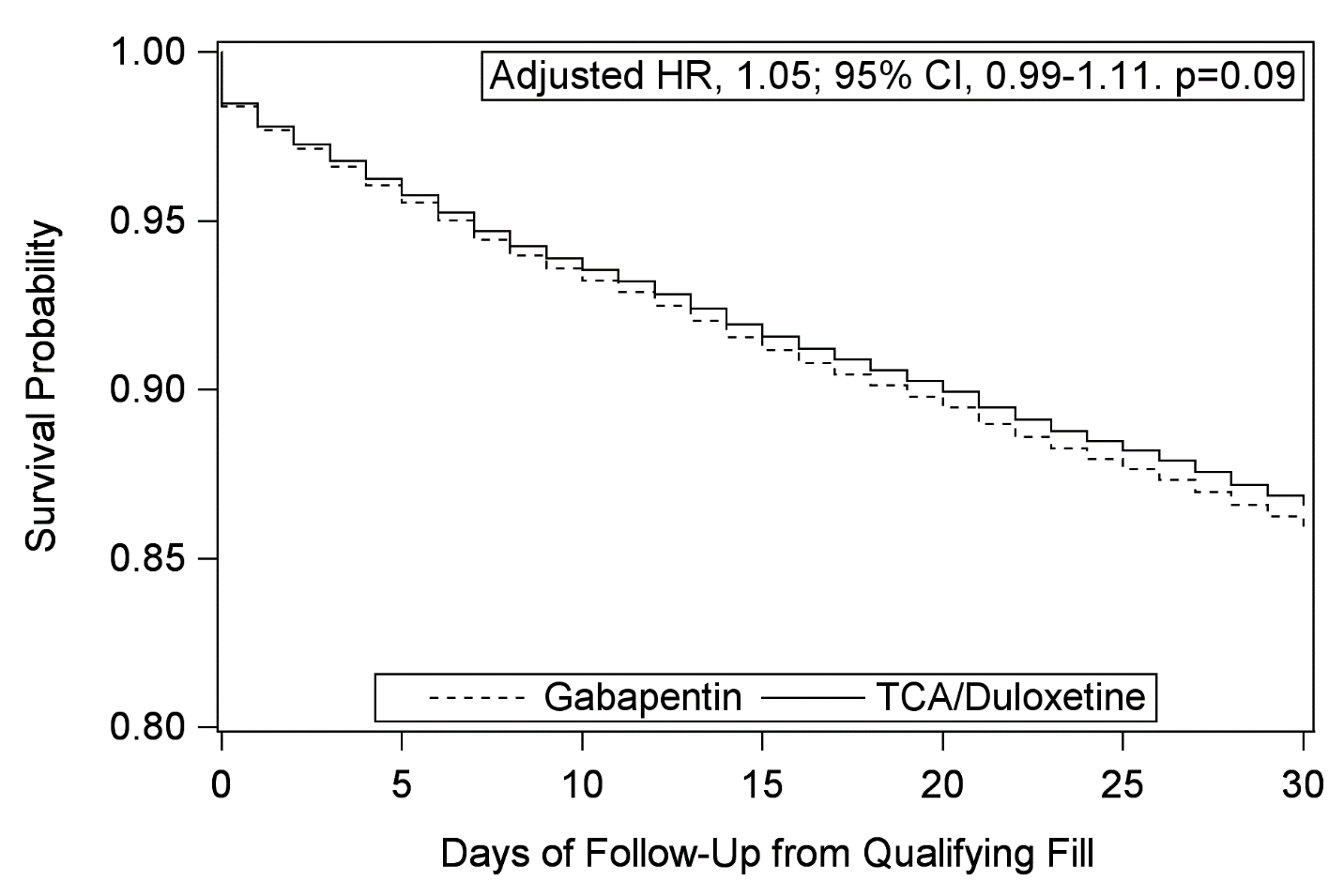
